# Learned Human Aortic Morphodynamics: A Dynamical-Systems Model of Post-EVAR Sac Remodeling

**DOI:** 10.1101/2025.09.29.25336910

**Authors:** Joseph A. Pugar, Junsung Kim, Michael Mansour, Charles Davis, Nhung Nguyen, Cheong Jun Lee, Trissa Babrowski, Hence Verhagen, Ross Milner, Andrei A. Klishin, Luka Pocivavsek

## Abstract

Biological tissue reorganizes in response to changes in its mechanical environment, yet quantitative models of that reorganization are almost always built where observations are dense. At organ-scale in living humans the observational data is sparse and noisy: a few irregularly spaced images per individual, acquired with variable instrumentation, with loss to follow-up and patient attrition. We ask whether governing equations for organ-scale remodeling can be recovered from this regime, and pose what those equations may reveal about the underlying biology.

We study the human abdominal aorta following endovascular aneurysm repair (EVAR), in which a stent graft excludes the aneurysm sac from arterial flow and thereby changes the mechanical loading of the sac wall. Each computed tomography (CT) scan is reduced to a two-dimensional state: surface area *Ã* (size) and fluctuation in integrated Gaussian curvature 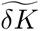 (shape), both normalized to a non-pathological aortic population. Across 100 patients and 220 scans pooled from two international centers, we use Z–SINDy, a sparse identification method with statistical-mechanical uncertainty quantification, to infer affine systems of ordinary differential equations (ODEs) governing 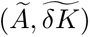 for anatomy that remodels successfully (regressing sacs) and anatomy that does not (stable sacs).

Both outcome classes approach stable fixed points, but at different locations in the morphological state space and on different timescales. Regressing anatomy approaches 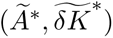 = (2.0,1.0) with characteristic timescales of 1.3 and 4.6 years; stable anatomy approaches its fixed point of (3.8, 2.5) with an order of magnitude slower timescale (10.9 and 16.6 years). Embedding the learned equations within Bayesian classifiers demonstrates that trajectory-based information resolves patient outcome classification earlier than positional evidence alone. Toward model utilization in future studies, these advantages are also observed when empirically observed loss to follow-up is carried through the priors. Recovering the equations that govern remodeling, rather than tracking its instantaneous state, therefore provides a foundation for clinical models built from the sparse records that routine imaging surveillance actually produces.

## 1 Introduction

Biological systems commonly exhibit continuous dynamical behaviors, yet quantitative models of their progression are typically built where observations are dense, controlled, and amenable to high-throughput acquisition. At the cellular scale, machine learning has inferred mechanistic models from images of cytoskeletal proteins [1] and swimming cells [2], inspired by the success of data-driven equation discovery for physical systems [3, 4]. At the anatomic scale, generative models trained on experimental movies recover noisy multicellular dynamics [5], and geometric frameworks extract deformation from whole-organ time-lapses during morphogenesis [6, 7]. The common thread is access to a temporally rich record of the process.

Organ-scale remodeling in living humans offers no such record. Measurements are irregular and sparse, often consisting of two to five images separated by months or years [8, 9], and carry segmentation variability and inter-scanner heterogeneity [10, 11]. Two problems separate this regime from the cellular and tissue-scale settings above. The first is identifying macroscopic latent variables that meaningfully capture organ remodeling patterns. The second is inferring low-order dynamics from time series that are short, noisy, irregularly sampled, and increasingly sparse over time. Both must be solved before such data can yield interpretable, generalizable equations of organ morphodynamics.

**Figure 1:**
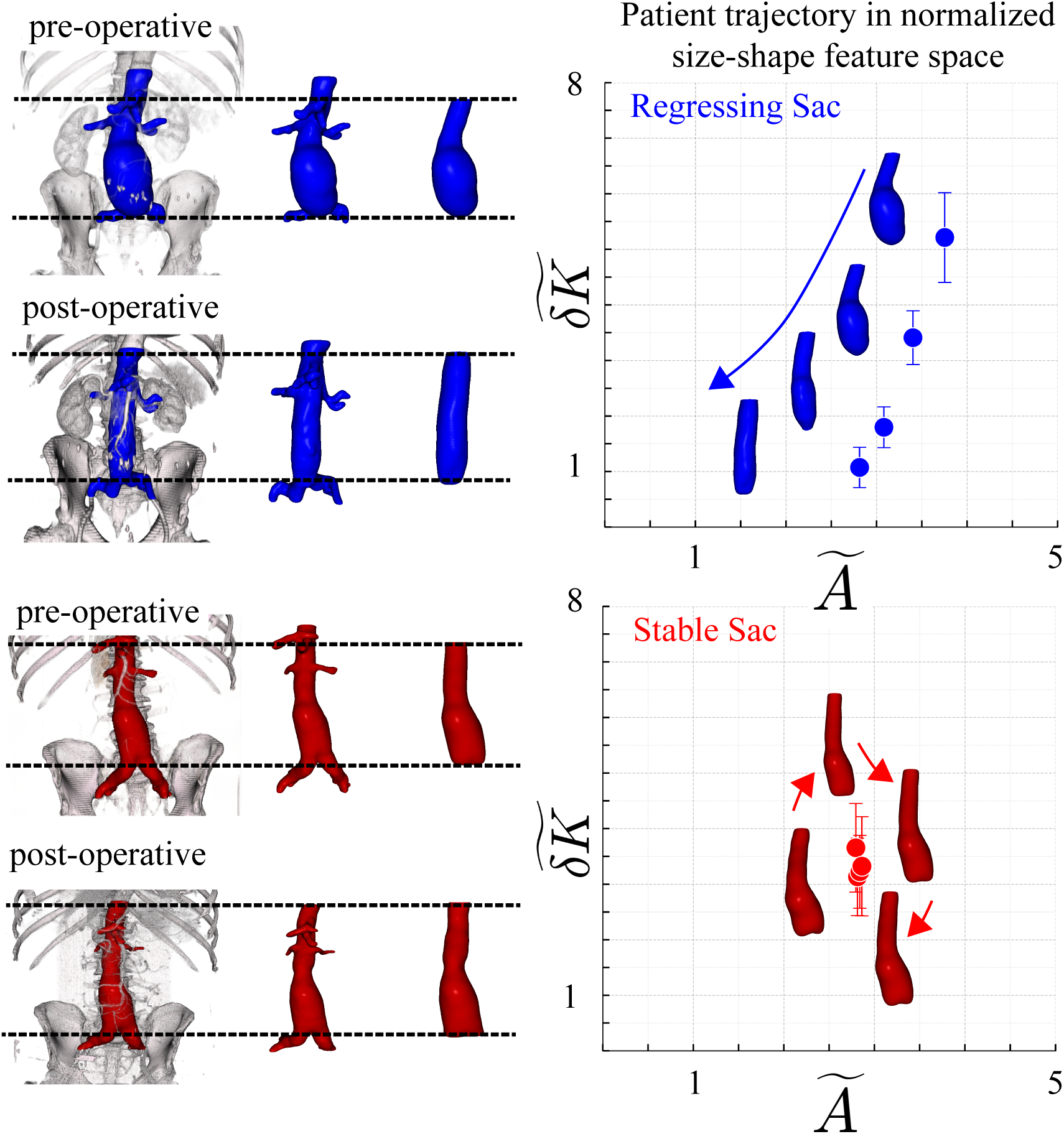
Defining aortic anatomy and mapping trajectories into size–shape space. Left: 3D segmentations of AAA scans before and after EVAR; dashed horizontal lines mark the superior and inferior cut planes used to standardize the analyzed sac segment across time points. Right: Each scan is projected into a normalized size–shape space (*Ã, δ̃K̃*), where *Ã* is normalized surface area and *δ̃K̃* the normalized fluctuation in integrated Gaussian curvature. Points are serial scans connected chronologically; arrows indicate temporal progression; error bars indicate the series of scales over which localized shape varies meaningfully [12]. Top row: regressing sac (blue; *>* 10%/year decrease in surface area). Bottom row: stable sac (red; *<* 10%/year change).

An abdominal aortic aneurysm (AAA) is a localized dilation of the aorta under pressurized flow. Endovascular aneurysm repair (EVAR) places a stent graft within the aorta to exclude the weakened aneurysm wall from continued stress [13, 14, 15]. This perturbation in the mechanical environment of the aorta is intended to induce favorable remodeling of the aneurysmal sac, and following the procedure, patients receive follow-up CT imaging of the aorta over years (Fig. 1). Postoperative surveillance and clinical decision-making are largely informed by discrete changes in aneurysm size that are extracted from longitudinal imaging [16, 17, 18].

Reducing an organ to a tractable state vector requires committing to macroscopic variables. Size alone (typically cross-sectional maximum diameter) is the conventional descriptor [19, 20, 21, 22], but it is insufficient as it fails to convey the geometric information of a deforming surface. Calibrated shape descriptors, in particular the fluctuation in integrated Gaussian curvature *δ̃K̃*, carry information complementary to size [23], and have been used to construct size–shape state spaces for aortic morphology [12]. We adopt the two-dimensional state **x** = [*Ã, δ̃K̃*]^T^, with each component normalized against a non-pathological aortic population so that the healthy morphology sits near (1, 1) and the coordinates are dimensionless. The compact size–shape latent space therefore provides a link between complex morphology and interpretable dynamics that can be learned from imaging data.

**Figure 2:**
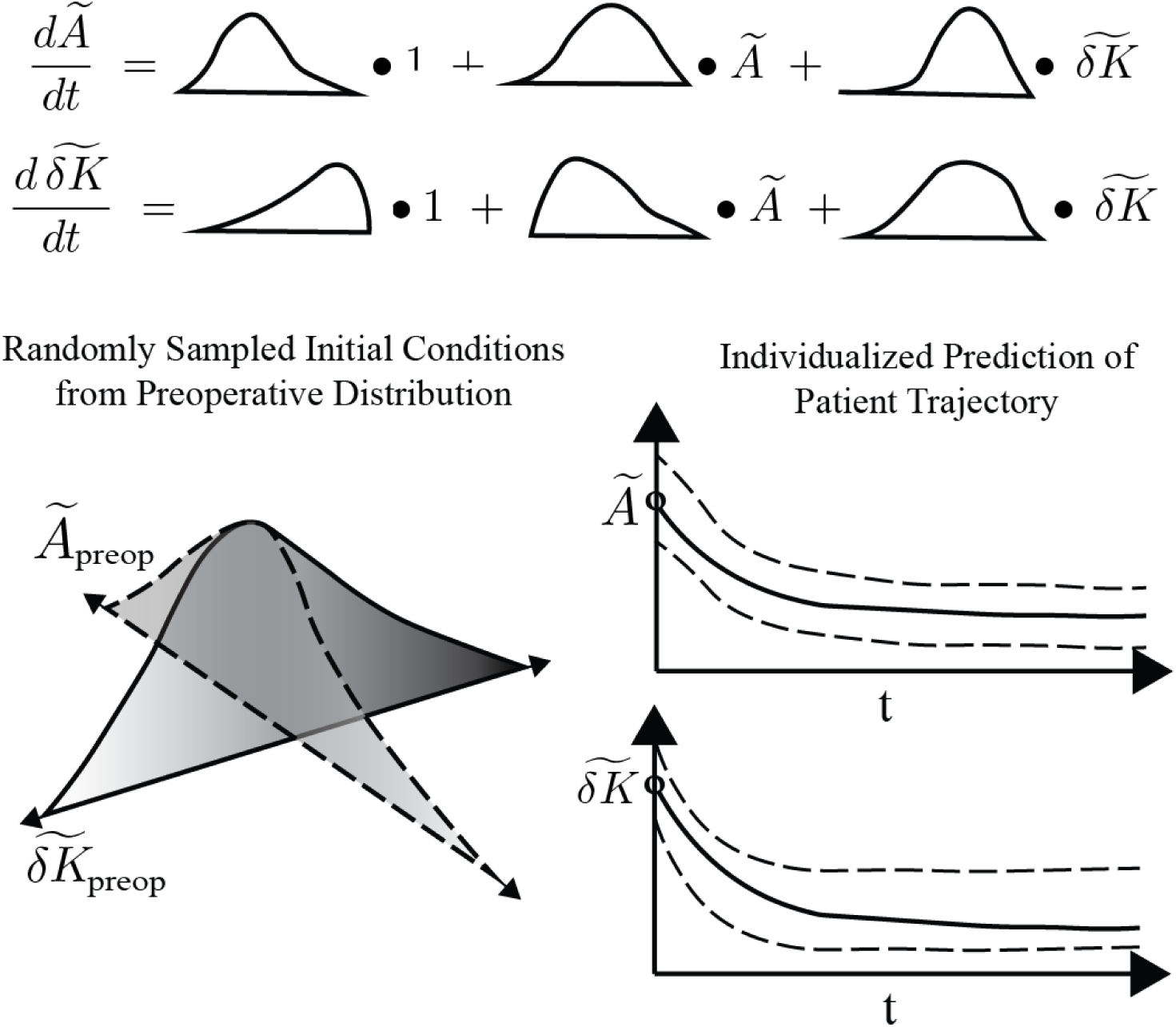
From posterior coefficients to individualized forecasts. Top: the learned system is affine in the two state coordinates, and Z–SINDy returns each of the six coefficients as a posterior distribution rather than a point estimate. Bottom left: initial conditions are drawn from the observed preoperative distribution of (*Ã_preop_, δ̃K̃_preop_*), so a simulated patient starts where real patients start. Bottom right: integrating a sampled coefficient set forward from a sampled initial condition yields a trajectory for each coordinate with a calibrated uncertainty band, which narrows as additional scans are incorporated.

A single CT scan cannot distinguish a sac that is regressing from one that is stable, because the two may occupy the same region of state space at any given moment. A coarse approximation of the relevant rate information is used in clinical practice, where remodeling is characterized by annual rates of change [24, 25]; this is supported by the observation that the failure of an aneurysm to regress carries worse long-term outcomes independent of other findings [16, 17, 26]. However, reducing the continuous evolution of remodeling behavior to single finite-difference rates of change over an approximated one-year interval discards nearly all of the temporal structure that could be learned when that information is used to fit a coupled ODE. Standard approaches to temporal modeling are also poorly matched to this regime. Linear mixed models describe each patient by a trend in time rather than by a flow in state space, and methods built for irregularly sampled series (e.g., Gaussian processes [27], latent ODEs [28], and neural controlled differential equations [29]) accommodate uneven observation times but either need far more data than a few hundred scans or return dynamics that are difficult to interpret [30, 31].

In this paper we recover governing equations for post-EVAR sac remodeling directly from clinical surveillance imaging, and we use those equations to forecast and classify individual patient trajectories. We use Z–SINDy [32], a sparse equation learning method that returns not only learned systems of ODEs from derivative training data but also posterior distributions over coefficients rather than point estimates [3, 4]. Sparsity of the equation functional form keeps the model interpretable at low data volume and is complementary to the parsimony of our size–shape reduced state space [33]. Because the inference returns a posterior, the fitted model is a generative object (Fig. 2): coefficient distributions can be resampled to create new trajectories within the learned dynamic variance. This places the model within the digital-twin paradigm, in which a virtual representation of a physical system simulates its behavior, is updated with data from its physical counterpart, and informs decisions [34, 35, 36]. In cardiovascular medicine the paradigm has developed around patient-specific simulation that combines mechanistic and statistical models [37, 38], and equation discovery sits between those two pillars (i.e., its equations are induced from data but integrate forward as a mechanistic model would). The result here is a low-dimensional, class-conditional digital twin of the postoperative aorta (one set of equations per outcome class, learned across patients and therefore closer to an aggregate than to an instance twin [34]), personalized through its initial condition and updated as each new scan revises a patient’s posterior class membership.

The paper proceeds in four steps that set the structure of the Results. First, we assemble and curate longitudinal CT records from two international centers (100 patients, 220 scans) and characterize the sparsity and attrition of the observational record itself. Second, we fit class-conditional affine ODEs to the regressing and stable cohorts and extract their fixed points, stability types, and relaxation timescales, showing that temporal upsampling by linear interpolation narrows the coefficient posteriors without changing the learned flow. Third, we integrate the posteriors forward to produce class-conditional ensembles of simulated patients, i.e., populations of digital twins carrying calibrated uncertainty bands. Fourth, we embed the learned equations in two Bayesian classifiers, one reading a patient’s position in state space and one reading their motion through it, and evaluate both on held-out patients, on the simulated ensembles, and under the empirically observed loss to follow-up.

## 2 Materials and Methods

### 2.1 Cohorts and morphological state

Longitudinal CT angiography was assembled from two centers (details in SI Section S1, Table S1). From each segmented aortic aneurysm we computed the surface area *Ã* and the fluctuation in integrated Gaussian curvature *δK* [23]. Both were normalized to the mean size and shape of a non-pathological reference population, yielding dimensionless *Ã* and *δ̃K̃* near (1, 1) for healthy aortas and increasing, in each dimension uniquely, with disease. The patient state is 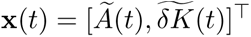. Construction of the non-pathological reference population and shape-scale averaging, resulting in the error bars in Fig. 1, follow our prior work [12]. Patients with clinical outcomes other than regressing or stable (e.g., endoleak requiring reintervention) were excluded from the analysis, but are discussed in the concluding remarks of the paper.

### 2.2 Curation of the raw scan record

Outlier rejection was applied to raw scans before any upsampling so that a rejected data point cannot seed an interpolated data point. Transitions were rejected on derivative magnitude using class-specific, anisotropic bounds: one-sided for regressing sacs, which decrease in size per clinical definition, and symmetric for stable sacs, which do not change substantially in either size or shape over a single year. Both endpoints of a flagged transition were removed from the dataset. Bound values, rationale, and sensitivity are given in SI Section S2.

### 2.3 Temporal upsampling and derivative estimation

Surviving scans were upsampled by linear interpolation onto a uniform time grid shared across patients, anchored at *t* = 0 (the time of repair; EVAR) and clipped per patient to their own observed range, with step Δ*t* = 1*/*12 yr. Interpolation is therefore never extrapolation, and no patient contributes evidence outside their own observation window. Derivatives were estimated by pairwise finite differences between successive observations within subject, anchored at the earlier endpoint throughout. Since the upsampled trajectories are piecewise linear, such a forward finite difference scheme induces no additional quadrature error over a centered finite difference.

To choose Δ*t*, we refit the model at a sequence of upsampling resolutions from the ground truth CT scans to daily spacing and tracked the fixed points and characteristic timescales of each class, quantities that, unlike the posterior width and variance explained, are not guaranteed to improve as interpolated rows are added. We selected Δ*t* = 1*/*12 yr as the canonical upsampling step, a coarse resolution at which the fixed points and timescales of both classes have stopped moving and agree with the values obtained from CT scans alone. The full resolution sweep and supporting diagnostics are given in SI Section S3.

### 2.4 Sparse identification (Z–SINDy)

We modeled the size–shape state space **x**(*t*) with Z–SINDy [32] using the affine library

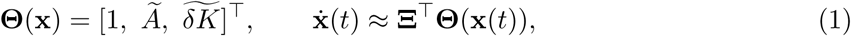

where the coefficients **Ξ** are inferred as a Bayesian posterior with resolution parameter *ρ* estimated from residual variance. Conditioning on a fixed active set gives a Gaussian posterior 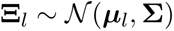 with ***µ*** = **C**^−1^**V** and **Σ** = *ρ*^2^**C**^−1^, where 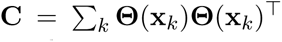 is library-library covariance, 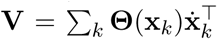 is the corresponding library-derivative covariance , and **Θ**(**x***_k_*) is the matrix of the library evaluated at every training row *k*. Both **C** and **V** are unnormalized and scale with the amount of interpolated data, affecting the reported uncertainty (SI Section S4). Z–SINDy selects an active set of parameters by minimizing a free energy *F* +*λn* that is penalized in proportion to the number of active terms *n*, with *λ* determining the strength of the regularization. Sweeping *λ* at the canonical upsampling step shows variance explained is maximized by the full six-coefficient system, with sparsification removing only coefficients whose posterior means are already near zero. We therefore fixed *λ* = −5 for all reported models so that both classes and both preprocessing pipelines are described by the same six-term, affine system. Likelihood, prior specification, and the sparsity sweep are given in SI Section S4 and Fig. S4.

### 2.5 Flow fields, fixed points, and ensembles

We calculated empirical and model dynamics in the 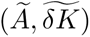 plane as per-patient difference vectors and mean Z–SINDy vector fields over a convex hull of the training data with fixed points and eigenvectors. Fixed points were obtained by solving 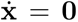, stability type was determined from the signs of the real parts of the Jacobian eigenvalues, and relaxation times from *τ* = 1*/*|Re *λ*|. Synthetic ensemble trajectories were generated by sampling 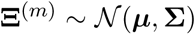 together with initial conditions from the observed preoperative distribution 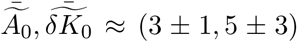 and integrating forward, with 1000 realizations per class.

### 2.6 Bayesian classification and validation

After the dynamic models were fit, two classifiers (one *static* in nature; one *dynamic* in nature) were evaluated on the same patients at the same time points. At time point *t* the static classifier uses a patient’s single most recent state at or before *t*, scored against time-indexed class ensembles, while the dynamic classifier uses every derivative estimate the patient has with 0 *< t_k_* ≤ *t*, scored against each class-conditional Z–SINDy model. A patient is evaluated at *t* only if their follow-up reached *t*. A decision is called confident when max*_c_ p*(*c* | ·) ≥ 0.9, accuracy is computed over that subset, and the fraction meeting the criterion is reported as classified. The 0.9 threshold is an arbitrary convention rather than a derived quantity, set to approximate the level of certainty at which a surgeon would act on a prediction rather than defer to the next surveillance scan. The convention also provides two unique metrics to evaluate model performance: 1. classification (*p >* 0.9): model decisiveness and 2. accuracy: performance once decisive. Likelihoods and normalization are in SI Section S5.

We performed four unique evaluations of the static and dynamic classifiers on the data. First, we performed in-sample classification: all of the available data was used to fit models and test them. Second, a leave-one-patient-out (LOO) method was used in which models were refit with a patient wholly excluded, so that every prediction comes from a model that has not seen the individual it is predicting. The third setting applies the two in-sample classifiers to the forward-integrated ensembles (synthetic trajectories) rather than to real patients, which removes the constraint that a subject can only be scored while their real follow-up continues. Individual-patient results appear in SI Section S6 and ensemble results in the main text, with Table S2 collecting all of them.

### 2.7 Attrition-aware priors

The fourth evaluation uses class-specific survival curves to compute changing priors at each subsequent classification timestep. Priors are therefore computed from the curated cohort as the fraction of each class still under surveillance at each postoperative time. In the previous three evaluations the class priors are fixed at *p* = 0.5 (equal regressing sac and stable sac probabilities) at all time points. In the attrition-aware analysis the class prior at time point *t* is set to the linearly interpolated surviving fraction, with baseline *p*(stable) = 0.53 taken from the curated cohort of 88 patients (47 stable and 41 regressing). Matched randomized dropout was applied to the simulated ensembles so that the number of trajectories contributing to each posterior declines on the same schedule. Results at one, two, and five years are given in the main text and at half-year resolution over the first eighteen months in SI Section S8.

## 3 Results

### 3.1 Cohort, curation, and the observational record

We assembled longitudinal CT angiography data sets from two centers: the University of Chicago (USA, *n* = 40 patients) and Erasmus University Medical Center (the Netherlands, *n* = 60), totaling 100 patients and 220 scans (Fig. 3A, Table S1). Among the 100, 44 were labeled regressing and 56 stable strictly using a size-only clinical metric of shrinkage ≥ 10%*/*year. Most patients contribute exactly two scans and none contribute more than four; the mean interval between consecutive scans is 1.9 years in the regressing group and 1.2 years in the stable group. The difference in follow-up cadence between centers is structural rather than incidental: Erasmus patients follow a protocol of one peri-operative and one 12-month scan, whereas Chicago patients have preoperative imaging at variable lead times and irregular follow-up extending to eight years (Fig. 3A). We treat the two centers as a single pooled dataset throughout where heterogeneity in scanner, cadence, and device are the main sources of noise.

**Figure 3:**
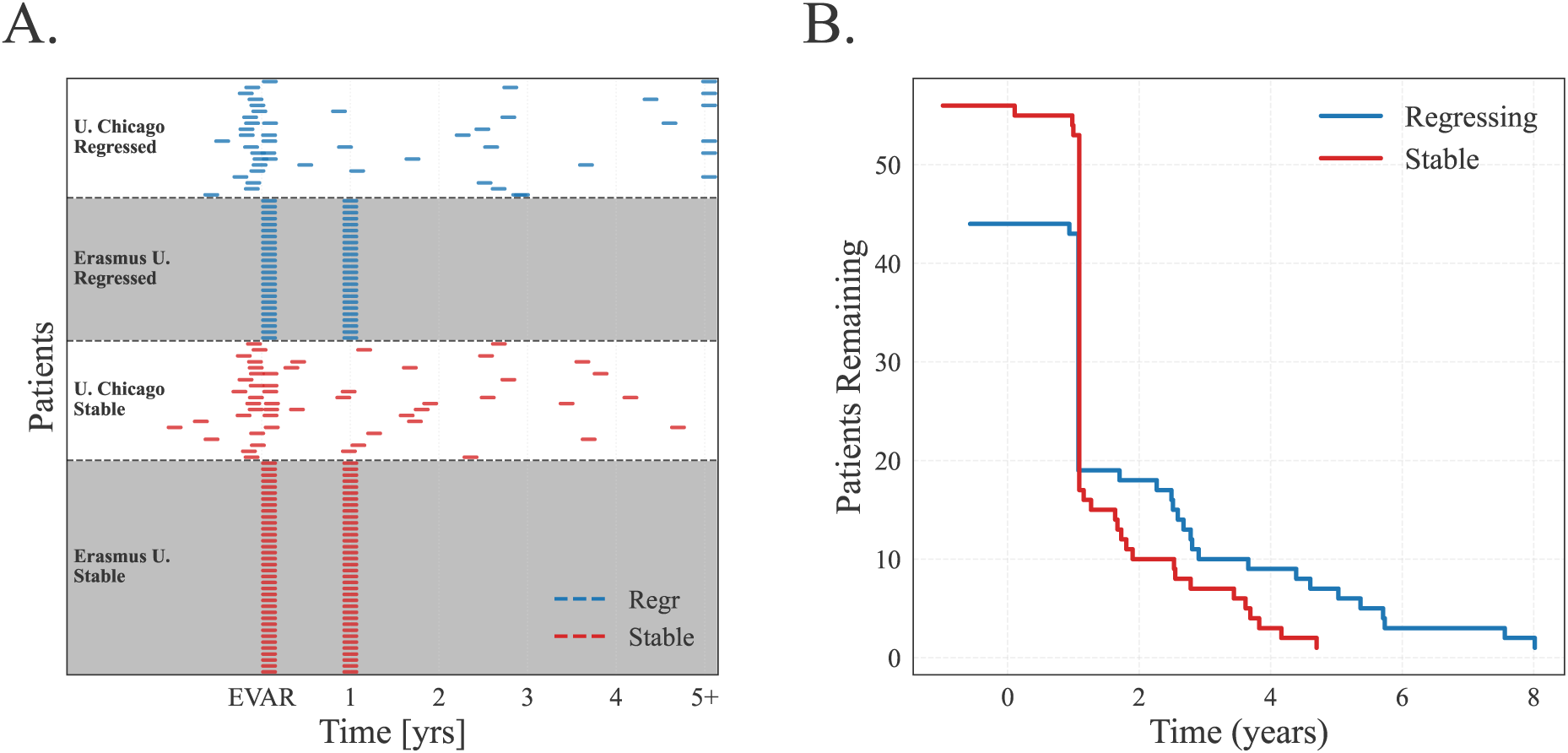
The observational record: schedule and attrition. (A) Per-patient scan timeline. Each row is a patient and each tick a CT scan, colored by outcome class and grouped by center. The Erasmus cohort’s protocolized two-scan schedule produces the dense vertical bands at repair and at twelve months, while the Chicago cohort supplies more erratic scans both preoperatively and through 5+ years postoperatively. (B) Number of patients in each class still under surveillance as a function of time since repair. Both classes are complete through the first year and contract sharply at approximately twelve months as the Erasmus protocol ends. Thereafter the stable class is lost faster than the regressing class, so the composition of the observed population drifts with time even though no individual patient changes.

Segmentation error and timestamp irregularity occasionally produce transitions between consecutive scans that are not physiologically plausible, and these are removed on the raw scans before any interpolation using a minimum-interval criterion and a class-specific bound on derivative magnitude (Section 2.2, SI Section S2). Of the 120 consecutive real-scan transitions entering this step, 14 were flagged and removed, 6 from the Chicago cohort and 8 from Erasmus. That the two centers lose comparable numbers from very different follow-up schedules is consistent with the flagged transitions reflecting segmentation errors rather than any property of the imaging protocol. After curation, 88 of the 100 total patients contribute at least one usable derivative.

Both attrition curves are essentially flat through the first year and then fall abruptly at approximately twelve months, where the Erasmus protocol ends and the cohort contracts to the Chicago patients: the regressing group drops from 44 to roughly 19 patients and the stable group from 56 to roughly 17. Beyond that point the stable curve declines faster, reaching its last observation before five years, while a small number of regressing patients remain under surveillance out to eight. By five years the surviving population is therefore both small and no longer representative of the enrolled one, being roughly six parts regressing to one part stable. Data attrition is a significant constraint to model fidelity at long time points, and we return to it in Section 3.6, where it is carried explicitly through the classifier priors to demonstrate how it can be better accounted for.

### 3.2 A parametric model of sac morphodynamics

We ask whether the coarse-grained dynamics of sac remodeling admit a low-order, interpretable description in the normalized size–shape space,

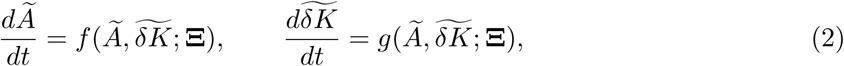

with 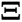 inferred over the affine library of Eq. (1). Sweeping the sparsity weight *λ* at the canonical upsampling step shows that variance explained is maximized by the full six-coefficient affine system, and that increasing *λ* removes terms only once it begins deleting coefficients whose posterior means are already near zero (Fig. S4, SI Section S4). All models reported below therefore retain the same six-term system.

What distinguishes Z–SINDy from a point-estimate fit is that each of those six coefficients is returned as a posterior distribution rather than a single number (Fig. 2). Two consequences follow, and they organize the remainder of the paper. First, the width of those distributions is itself a quantity to be managed: it determines whether the fitted equations can be integrated forward with physiologically reasonable bounds. Second, once the widths are narrow enough, the equations become usefully generative. Drawing a coefficient set from the posterior and an initial condition from the observed preoperative distribution and integrating produces a single simulated patient, and repeating produces a population of them, collectively carrying a calibrated band rather than a point prediction. We refer to a model used this way as a low-dimensional digital twin: a forward-integrable simulator of an individual aorta, conditioned on outcome class, personalized through its initial condition, and refinable as further scans arrive.

Both consequences depend on how derivatives are estimated from the sparse record. Interpolated rows enter the posterior covariance exactly as additional independent observations would, so the posterior width shrinks as Δ*t* → 0 whether or not the added rows carry new information, and cannot by itself select the upsampling step. We therefore selected Δ*t* = 1*/*12 yr as the canonical upsampling timestep, from quantities that are not guaranteed to improve by construction, the fixed points and characteristic timescales of each class, which stabilize below Δ*t* ≈ 0.5 yr and agree with the values obtained from the CT scans alone (Methods, SI Section S3).

**Table 1:** Learned coefficients of the affine Z–SINDy models. Entries are posterior mean ± one standard deviation. Standard deviations are colored in red if the resulting coefficient distribution they form straddles zero. The constant term sets the location of the fixed point and the *Ã* and *δ̃K̃* terms set the relaxation rates. Fixed points, eigenvalues, and relaxation times derived from these coefficients are given in Table 2. The same posterior standard deviations are shown graphically, across the full range of upsampling steps, in Fig. S2.

| | 1 | $\tilde{A}$ | $\tilde{\delta K}$ |
| --- | --- | --- | --- |
| **CT scans** |  |  |  |
| *Regressing* |  |  |  |
| $\tilde{A}$ | 0.611 ± **0.661** | $-0.328 \pm 0.211$ | $0.011 \pm$ **0.066** |
| $\tilde{\delta K}$ | $-0.307 \pm$ **0.661** | $0.578 \pm 0.211$ | $-0.799 \pm 0.066$ |
| *Stable* |  |  |  |
| $\tilde{A}$ | $0.382 \pm 0.353$ | $-0.104 \pm$ **0.119** | $-0.002 \pm$ **0.034** |
| $\tilde{\delta K}$ | $-0.087 \pm$ **0.353** | $0.160 \pm 0.119$ | $-0.152 \pm 0.034$ |
| **Upsampled, $\Delta t = 1/12$ yr** |  |  |  |
| *Regressing* |  |  |  |
| $\tilde{A}$ | $0.418 \pm 0.149$ | $-0.200 \pm 0.058$ | $-0.019 \pm$ **0.022** |
| $\tilde{\delta K}$ | $-0.158 \pm 0.149$ | $0.462 \pm 0.058$ | $-0.768 \pm 0.022$ |
| *Stable* |  |  |  |
| $\tilde{A}$ | $0.196 \pm 0.086$ | $-0.048 \pm 0.030$ | $-0.006 \pm$ **0.008** |
| $\tilde{\delta K}$ | $-0.137 \pm 0.086$ | $0.103 \pm 0.030$ | $-0.105 \pm 0.008$ |

### 3.3 Learned flows, fixed points, and timescales

We fit two Z–SINDy models: one to the CT dataset, comprising 46 regressing and 55 stable derivatives, and one to the upsampled trajectories, comprising 1070 and 861. The means of the two models are shown side by side in Fig. 4 and their coefficients including uncertainty in Table 1. The fitted models share a common structure across both classes and both preprocessing pipelines, as shown by the fixed point and timescale agreement in Table 2. In every case, the Jacobian has two real, negative eigenvalues, so each class is governed by a stable node. Regressing and stable sacs are therefore distinguished not by the type of dynamics but by the location of the fixed point and the rate of approach to it.

Regressing sacs flow toward (2.0, 1.0), where the shape coordinate has returned to approximately the non-pathological reference value of 1 while the size coordinate remains at roughly twice the reference surface area of a healthy abdominal aorta. Stable sacs flow toward (3.8, 2.5), with both coordinates elevated relative to healthy aortas. The two targets are separated in both coordinates, by a factor of roughly 1.9 in size and 2.5 in shape.

**Figure 4:**
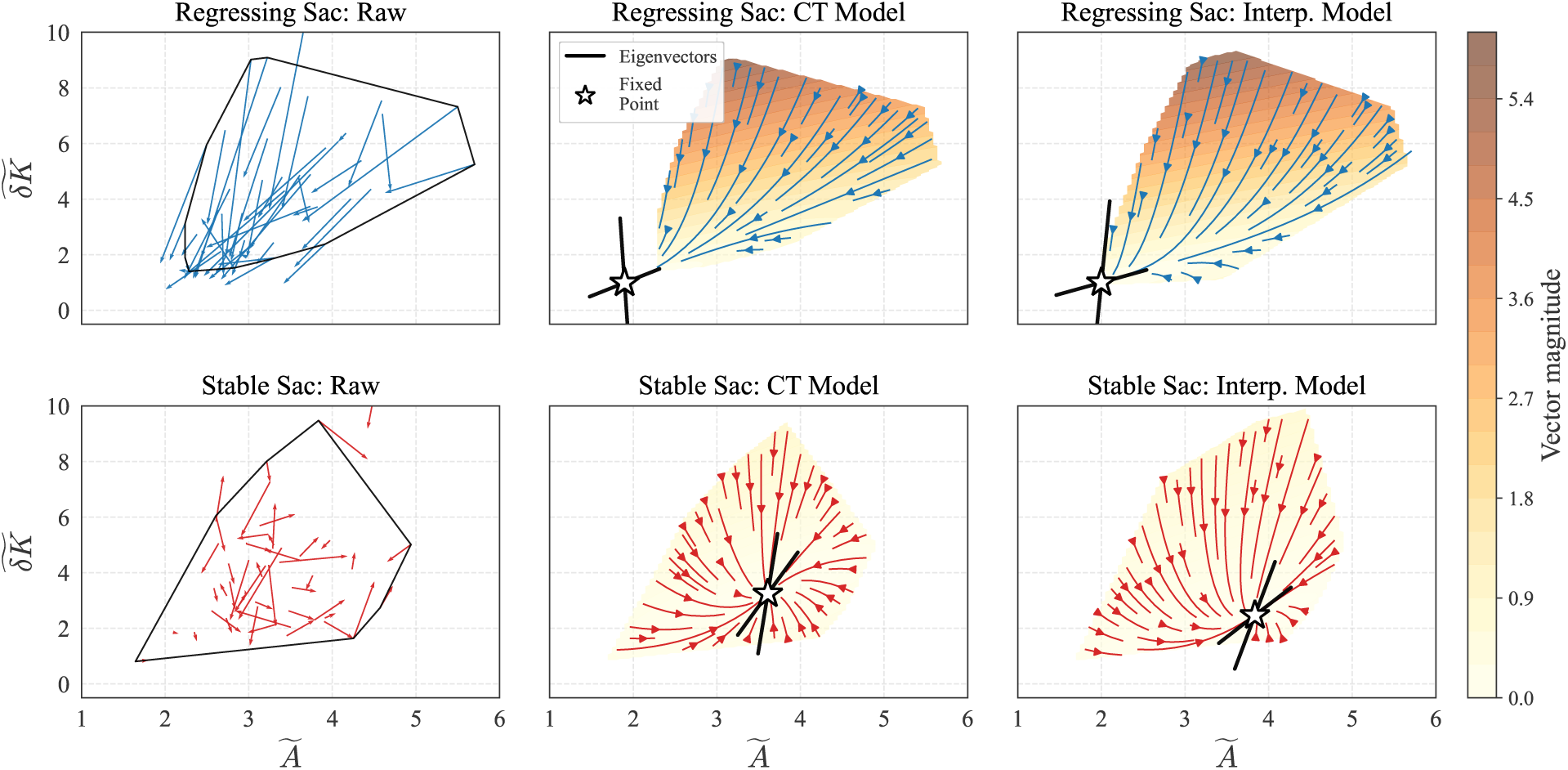
The learned dynamical systems have similar properties with and without interpolation. Rows: regressing sacs (top, blue) and stable sacs (bottom, red). Left column: per-patient finite-difference vectors on the curated raw scans, with the convex hull of observed states outlined. Center column: the mean Z–SINDy vector field fit to CT scans only. Right column: the mean Z–SINDy vector field fit to the canonical upsampled trajectories at Δ*t* = 1*/*12 yr. Stars mark class-specific fixed points, black lines the local eigenvectors, and background shading the vector magnitude on a shared scale. The two model columns agree in fixed-point location, stability type (a stable node in both classes), and the orientation of the principal directions. Regressing sacs carry large vector magnitudes directed down and to the left, toward a fixed point at the lower-left corner of the observed region, whereas stable sacs move slowly toward a fixed point sitting inside the region they already occupy.

The eigenvalues of the regressing model, −0.22 and −0.75 yr^−1^, correspond to relaxation times of 4.6 and 1.3 years. The eigenvector of the faster mode lies almost along the shape axis, so shape normalizes over roughly 1.3 years while the mixed size–shape mode decays over roughly 4.6 years. The stable model approaches its own fixed point on timescales of 16.6 and 10.9 years, longer than any realistic surveillance window and without a comparable separation between the two modes.

The CT models place their fixed points at (1.9, 1.0) and (3.6, 3.2) and return the same stability type and the same ordering of timescales, with the regressing class faster than the stable class by a factor of roughly three. What the two pipelines do not share is the magnitude of uncertainty. As expected, the CT models have considerably wider coefficient distributions than those of the upsampled models.

Variance explained against the raw scans (Fig. S3) is modest in absolute terms (0.12 and 0.10 for the CT-only models, 0.12 and 0.06 after upsampling), which is expected for a model of this form. An affine flow in two coordinates is not intended to predict individual scan-to-scan transitions, but rather to describe the mean field about which those transitions scatter. Support for that mean field comes instead from its reproducibility across two independent preprocessing pipelines and across the interpolation sweep, and from its performance on patients excluded from fitting, reported below.

**Table 2:** Fixed points, eigenvalues, and relaxation times for both classes under both preprocessing pipelines. Fixed points and eigenvalues are computed from the unregularized posterior mean; *τ* = 1*/*|Re *λ*|. Every eigenvalue is real and negative: all four fits approach stable nodes. The dependence of these four quantities on the upsampling step is shown in Fig. S1.

| Dataset | Class | $\tilde{A}^*$ | $\tilde{\delta K}^*$ | $\lambda_{1,2}$ (yr$^{-1}$) | $\tau_{0,1}$ (yr) |
| --- | --- | --- | --- | --- | --- |
| CT | Regressing | 1.90 | 0.99 | $-0.315, -0.812$ | 3.2, 1.2 |
| CT | Stable | 3.61 | 3.24 | $-0.113, -0.143$ | 8.9, 7.0 |
| Upsampled | Regressing | 2.00 | 1.00 | $-0.216, -0.752$ | 4.6, 1.3 |
| Upsampled | Stable | 3.84 | 2.47 | $-0.060, -0.092$ | 16.6, 10.9 |

### 3.4 Forward-integrated forecasts carry calibrated uncertainty

The practical consequence of the coefficient posteriors is realized by integrating them forward. Table 2 compares the CT and interpolated models through their fixed points, eigenvalues, and relaxation times; Fig. 5 compares them in the way the models are used, i.e., as forecasting simulators. For each pipeline and each class we drew 1000 coefficient sets from the posterior together with initial conditions from the same observed preoperative distribution 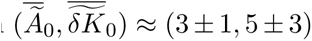, so the only difference between the two ensembles is the coefficient posterior itself.

The mean trajectories are close in all four panels, which is the temporal counterpart of the fixed-point agreement in Table 2. They are nearly coincident over the first two years, the interval in which patients are still reliably imaged, so the two pipelines produce the same forecast over the horizon the record actually covers, and they diverge only modestly thereafter. The band widths do not agree. By five years the CT ensembles span most of the observed region of state space and extend into morphological descriptions that are not interpretable, including *δ̃K̃* below zero in the regressing class, i.e., a shape fluctuation of the wrong sign. This follows directly from the coefficient posteriors of Table 1. Several terms in the CT models have posterior standard deviations comparable to their means, so a non-negligible fraction of draws carries at least one coefficient of the wrong sign, and draws in which a self-coupling term flips positive grow without bound. Those divergent realizations fill the CT band.

This figure should not be read as showing that the interpolated posterior is calibrated in the statistical sense (i.e., that a nominal 90% band contains 90% of held-out observations). Selecting Δ*t* fixes the posterior mean, whose fixed points and timescales stop moving within the plateau, but not the posterior width, which keeps contracting with the number of interpolated rows across the entire plateau whether or not those rows are informative (SI Section S3, Fig. S2). What the figure does establish is separate from that concern. The mean flow is the same object under both pipelines over the horizon the data support, and the CT posterior admits sign changes in several coefficients, so ensembles drawn from it cannot be integrated forward as forecasts even where the CT point estimates are sound. Forward integration, and everything built on it below, is practical here only because the interpolation step narrowed the coefficient posteriors sufficiently for the resulting trajectory bands to remain within interpretable regions of the size–shape space over a five-year horizon.

**Figure 5:**
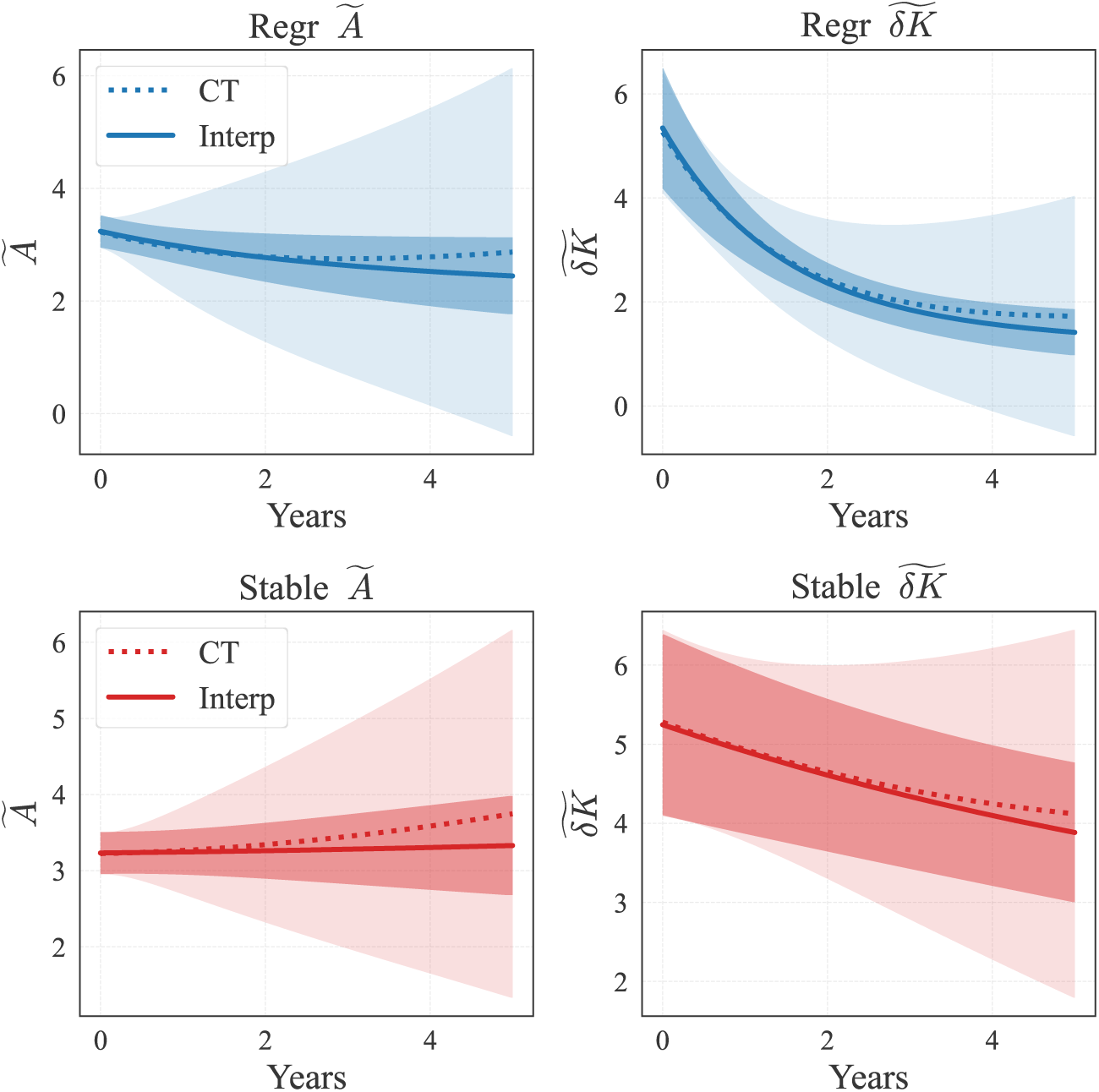
Forward-integrated ensembles from the CT and interpolated models. Rows: regressing (top, blue) and stable (bottom, red). Columns: *Ã* and *δ̃K̃*. Solid lines give the ensemble mean of the interpolated model and dotted lines that of the CT model, with the dark band giving the interpolated spread and the light band the CT spread. Both ensembles use 1000 realizations per class and draw initial conditions from the same observed preoperative distribution, so the difference between the bands reflects the coefficient posteriors alone. The means track each other closely through the first two years, while the CT band widens into regions of state space that are not physically admissible.

### 3.5 Trajectory evidence separates the classes within a year

A patient’s trajectory through the size–shape state space carries information beyond the instantaneous coordinates. We tested this by embedding the class-conditional models in two Bayesian classifiers, one reading position (static classifier) and one reading motion (dynamic classifier). The static classifier scores a patient’s most recent state (*Ã, δ̃K̃*) against time-indexed class ensembles, while the dynamic classifier scores every derivative estimate accumulated up to that time against each class-conditional ODE. A decision is recorded as confident when the returned posterior falls outside the band [0.1, 0.9], i.e., a labeled prediction is not made unless it carries *>* 90% confidence (Methods).

The cleanest arena for comparing the two rules is the simulated ensembles, because the observed cohort cannot answer how early the two regimes would separate under regular follow-up: no patient is imaged regularly and few are imaged for long. Sampling coefficients from the posterior together with initial conditions from the observed preoperative distribution (*Ã_preop_, δ̃K̃_preop_*) and integrating forward yields 1000 trajectories per class (Fig. 6, left), each a simulated patient in the sense of Fig. 2.

The ensembles reproduce the structure of the fitted flows and make its timing explicit. Immediately after repair the two classes occupy the same region: the marginal densities of *Ã* and δ̃K̃ are coincident and the joint densities overlap completely. Separation appears first in shape and then in size, following the timescale ordering in Table 2: by one year the *δ̃K̃* marginals are clearly displaced while the *Ã* marginals still overlap substantially, and by two years both coordinates have separated. By five years the regressing ensemble has collapsed onto a narrow band of low shape values while remaining broad in size, and the stable ensemble sits higher in both coordinates with much the wider shape distribution, so the two classes differ in spread as much as in location.

Applying the two classifiers to the same trajectories makes the value of motion explicit (Fig. 6, right). The dynamic classifier resolves roughly four in five subjects by one year, a classification fraction that the static classifier does not match even at five. The static classifier improves steadily as the ensembles drift apart, which is the expected behavior of a positional readout, but it has resolved only a little over half of the subjects by the point where real surveillance has all but ended.

Because the trajectories are generated by the same models being evaluated, this analysis characterizes the information content of the learned flows rather than validating them against independent data. It provides an upper bound on the separability achievable by any classifier restricted to these two coordinates under clean, regular sampling. The gap between that bound and the individual-patient results of the next section is therefore a measure of what the observational record costs, and it is attributable to scan timing, segmentation error, and loss to follow-up rather than to the equations.

**Figure 6:**
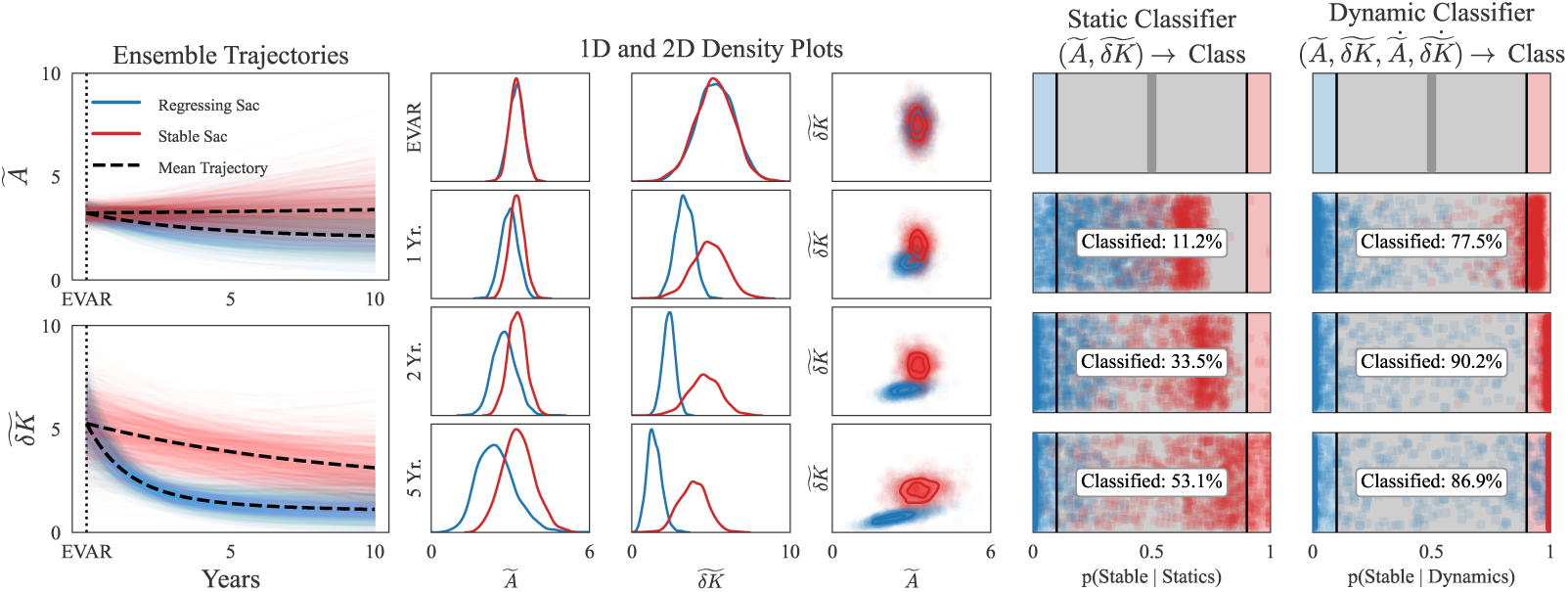
Class-conditional ensembles and their separability under ideal follow-up. Left: 1000 forward-integrated trajectories per class in each coordinate, sampling both the coefficient posterior and the observed preoperative initial-condition distribution; dashed lines are ensemble means. Center: marginal densities of *Ã* and *δ̃K̃* and their joint density at repair and at one, two, and five years. The classes are coincident at repair and separate first in *δ̃K̃*, consistent with the shape-aligned fast eigenmode. Right: posterior probabilities returned by the static and dynamic classifiers on the same trajectories, with the fraction leaving the uncertain band [0.1, 0.9] annotated. The dynamic classifier resolves 79.6% of subjects by one year, which the static classifier does not match by five.

### 3.6 Classification under empirically observed loss to follow-up

The same ordering holds when the two classifiers are evaluated on the real observational record, both in-sample and leave-one-patient-out (SI Section S6): the dynamic classifier commits on more than nine in ten eligible patients at every time point while the static classifier commits on fewer than one in ten, and the leave-one-patient-out results establish that the learned flows generalize to patient derivatives not used in model fitting. What those experiments cannot do is characterize the classifiers beyond eighteen months, because sparsity and attrition thin the eligible cohort to a third of its enrolled size by then (i.e., the record runs out before the dynamics do). The simulated ensembles have no such horizon, but using them at long times raises a problem the fixed-prior evaluations ignore: they treat the classified population as the enrolled population, which the survival curve does not support (Fig. 3B). If stable sacs are lost more rapidly than regressing ones, then the probability that a subject still under observation at five years belongs to the stable class is not the enrollment proportion, and a classifier using the enrollment prior is misspecified in a known direction. We therefore re-evaluated both classifiers with the class prior at each time point set to the empirically observed surviving fraction, and with matched randomized dropout applied to the simulated ensembles so that the number of trajectories contributing to each posterior declines on the same schedule as the real cohort (Fig. 7). Of the 2000 simulated subjects, 1933 remain at one year, 587 at two, and 168 at five, which reproduces the real schedule over the interval where the two can be compared (85 of 88 patients at one year).

In this attrition-aware classification analysis, the prior moves substantially over the 5-year experiment. It begins at *p*(stable) = 0.53 against *p*(regressing) = 0.47, is unchanged at one year because both classes are still essentially complete, and then shifts to 0.39 against 0.61 at two years and 0.14 against 0.86 at five, as stable patients are preferentially lost.

Under these conditions both rules become reliable once they commit, exceeding 95% accuracy on the subjects they commit to, but they commit on very different schedules (Fig. 7). The trajectory-based rule resolves four subjects in five within the first year, while the position-based rule resolves one in eight at the same point and does not exceed two-thirds until two years, by which time roughly a third of the enrolled cohort is still under surveillance. Resolving the same question at half-year resolution over the first eighteen months gives the same ordering (SI Section S8, Fig. S8).

Two contributions to this result should be distinguished. That performance does not degrade under a time-dependent prior reflects in part a property of the fitted flows, since the regressing ensemble converges toward a compact region of state space and is therefore more predictable than the diffuse stable ensemble, so up-weighting the regressing class as follow-up proceeds carries a smaller information penalty than it otherwise would. It also reflects the fact that the analysis was performed on model-generated trajectories subject to imposed dropout rather than on the surviving patients themselves, which the cohort is too small to support at five years. These results therefore characterize the behavior of the inference under realistically thinning observation, and do not constitute an independent test of the learned flows. What they establish is narrower and still useful: the interval over which a trajectory-based classification becomes decisive falls inside the interval over which patients are actually being imaged, whereas the interval over which a positional-based classification becomes decisive does not.

**Figure 7:**
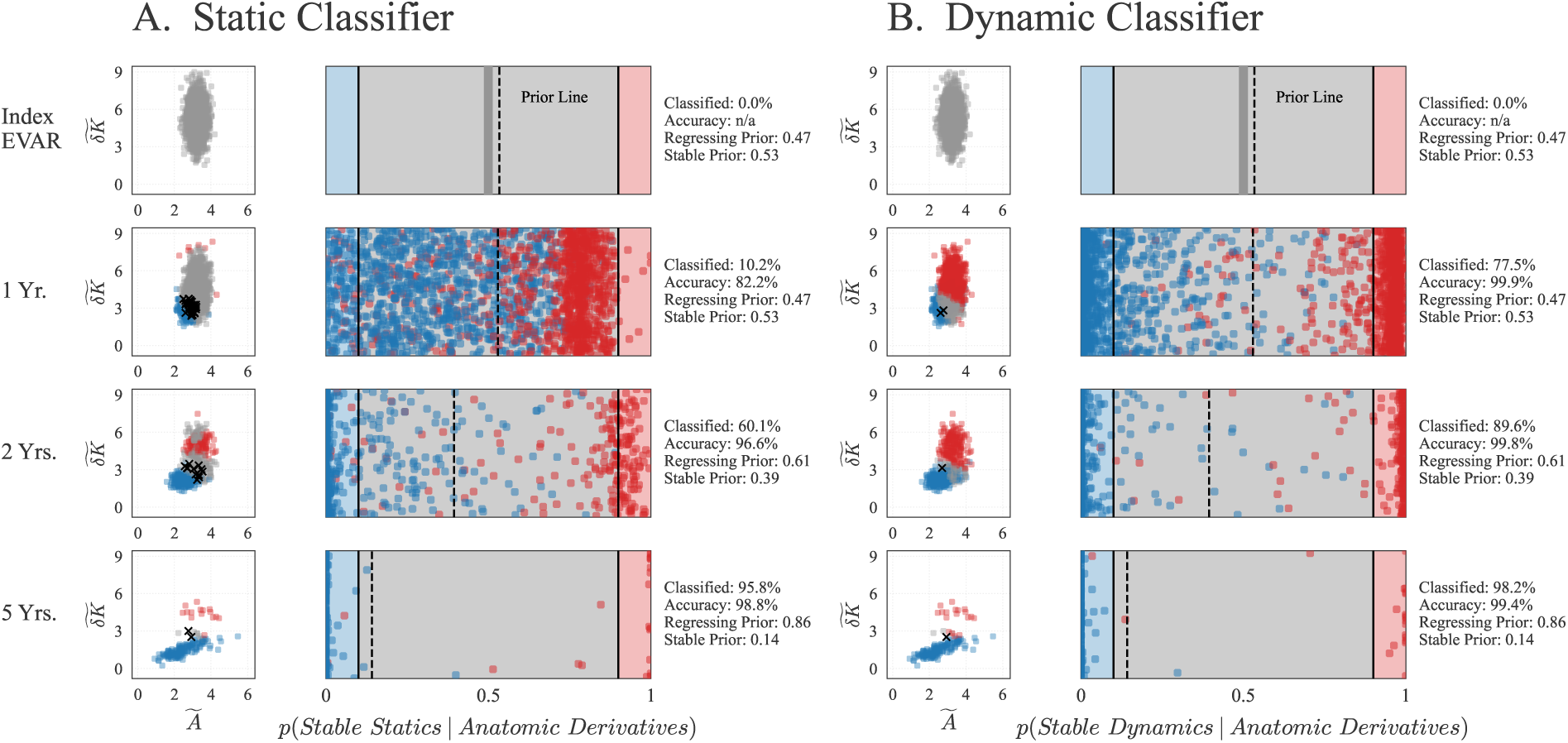
Classification under empirically observed loss to follow-up. (A) Static classifier. (B) Dynamic classifier. Within each block the left column gives the posterior *p*(stable | ·) for every simulated subject (blue regressing, red stable), with the uncertain band [0.1, 0.9] shaded and the class prior at that time point marked by the dashed line, and the right column gives the corresponding instantaneous (*Ã, δ̃K̃*) distribution; accurate classifications in their respective colors, unclassified in grey, and misclassified as black x’s. Rows are index EVAR and one, two, and five years. Class priors are set to the surviving fraction measured in the real cohort (Fig. 3B), moving from 0.47*/*0.53 regressing to stable at repair to 0.86*/*0.14 at five years, and matched randomized dropout is applied to the ensembles so that the number of contributing trajectories declines on the same schedule. The dynamic classifier resolves 77.5% of subjects at one year against 10.2% for the static classifier, and both exceed 80% accuracy on the subjects they commit to.

## 4 Discussion

In this paper we show that affine differential equations governing organ-scale remodeling are recoverable from typical clinical CT surveillance. Reducing the aorta to two macroscopic geometric coordinates and applying sparse equation discovery with uncertainty quantification yields coefficients that are directly interpretable, where the constant terms locate the morphology toward which a sac moves and the linear terms set the rate of approach to it. Because the inference returns posteriors rather than point estimates, those coefficients also define a mean-field, per-cohort and per-outcome probabilistic digital twin of the postoperative sac, and it is that object, rather than any single fitted curve, that the remainder of this section interprets.

### 4.1 Learned morphodynamics of the post-EVAR sac

Regressing and stable sacs are both governed by a stable node. Failure to regress is therefore not a distinct dynamical regime but the same relaxation toward a different target with a time constant roughly eight times longer, which over a five-year surveillance window is difficult to distinguish from stasis. The distinction is mechanistically consequential, since there is growing discussion of whether stable sacs post-EVAR are associated with worse outcomes or should be considered suboptimal results [16, 39, 40].

The regressing fixed point sits at a shape coordinate near the non-pathological reference value and a size coordinate at roughly twice that reference, so mechanical unloading of the sac wall restores surface regularity before restoring size, with an eigenvector lying almost along *δ̃K̃* and a relaxation time of 1.3 years against 4.6 years for the mixed mode. The geometric signature of successful remodeling therefore precedes the size change that current surveillance is designed to detect, which is consistent with the observation that most sac regression occurs within two years of repair [41].

### 4.2 The role of temporal upsampling

Linear interpolation imposes a smoothness assumption between consecutive scans of the same patient and adds no information, since the densified series is a deterministic function of the measured one which comes directly from the sparse CT data. What it changes is the uncertainty of the models. Derivatives estimated on a common grid are comparable across patients with different follow-up schedules; the model is trained using the interior of the state space rather than the endpoints of sparse vectors; and the resulting posterior is narrow enough to reliably integrate forward many times over fluctuating initial conditions (preoperative states). Three results bound the assumption. (1) The fixed points and both timescales stop moving below Δ*t* ≈ 0.5 yr, so nothing reported here depends on how finely we interpolate within the plateau. (2) The same pipeline applied to CT scans recovers the same stability type and fixed points agreeing to within the width of the posteriors, so the flow is a property of the measurements rather than of the upsampling step. (3) Coarse interpolation degrades every diagnostic, which places the failure mode where it should be, at grids coarser than the data they resample.

### 4.3 Bayesian classifiers

The two classifiers test the learned models, and the three settings in which they were evaluated answer three distinct questions. (1) Do the learned flows carry class information on real, CT-only clinical follow-up? On the observed cohort, evaluated leave-one-patient-out so that no prediction comes from a model that has seen the individual, positional evidence rarely reaches confidence, which is the quantitative form of the clinical observation that a single postoperative scan does not indicate what a sac will do, while trajectory evidence resolves nearly every patient. (2) How early could these two coordinates resolve the classes under regular follow-up? On the synthetic ensembles, which bound what any classifier restricted to this state space could achieve, the bound is reached within a year by the trajectory readout and not within five by the static positioning. (3) How does the inference behave when the population being classified thins the way real populations thin? Under attrition-matched dropout, the ordering is unchanged. The consistent finding across all three settings is that consideration of where a sac is heading resolves its class earlier, and for more subjects, than consideration of where it is.

Loss to follow-up was accounted for through the class priors rather than ignored. Surveillance thins steeply and unequally between the classes, and by five years the surviving population is roughly six parts regressing to one part stable, so the probability that an observed subject is stable differs from the enrollment proportion by a factor of nearly four. Re-evaluating both classifiers under time-dependent priors and matched dropout leaves the ordering between them unchanged and does not degrade accuracy, though the static rule commits more often at late times because by then the prior itself has become strongly informative (Table S2). Because that analysis uses model-generated trajectories with imposed dropout rather than the surviving patients, which the cohort cannot support at five years, it describes the behavior of the inference under thinning observation and is not an independent test of the dynamics.

### 4.4 Limitations

Several features limit how far these results extend. Most patients contribute exactly two scans and therefore one measured finite difference, so they constrain the flow along a straight segment rather than an observed trajectory and therefore a single patient cannot inform ordered dynamics. Rather, the ensemble of linear trajectories identifies mean-field dynamical behavior that is ordered, and produces a mean-field, per-cohort and per-outcome probabilistic digital twin rather than a per-patient one. This is also the reason the dynamic classifier is decisive but imperfect on real patients: a sharp posterior built from a single measured vector is confident without being especially well informed. Curation is class-aware by construction (SI Section S2). Eligibility at each time point requires follow-up to have reached it, so the 1.5-year results describe 29 of 88 patients with atypically long surveillance. The state space is restricted to *A* and *δ̃K̃* to increase interpretability, but thrombus morphology, graft configuration, and hemodynamic variables may resolve structure a two-dimensional state cannot, as may more principled shape metrics from differential geometry [42] in place of the empirically motivated *δK* [23]. Validation is leave-one-patient-out within a pooled two-center dataset, so external validation and a temporal extrapolation test remain to be done.

### 4.5 Future work

The gap between the ensemble bound and the held-out patient results measures what the current observational record costs, and it defines what the next study should be. A surveillance protocol with imaging at regular intervals through the first two postoperative years would supply each patient with several measured derivatives instead of one, closing most of the distance between the real-patient and ensemble results. Within the existing retrospective setting, the natural extensions are external validation at more centers, a temporal extrapolation test in which models fit on early follow-up predict late follow-up, and a third dynamical class for failing anatomy such as endoleak, which the present two-class framework excludes by design. Each of these additional batches of data sharpens the same probabilistic digital twin model. Regular early imaging would narrow the coefficient posteriors of the mean-field, per-cohort and per-outcome probabilistic digital twin using measured rather than interpolated derivatives, external cohorts would establish whether one set of class-conditional equations transfers across centers or whether each cohort carries its own twin, and a third dynamical class would add a failing-anatomy twin alongside the regressing and stable ones.

The approach also generalizes beyond this organ and this intervention. One defines a low-dimensional macroscopic state from segmentation output, learns class-conditional equations with calibrated uncertainty, and uses the resulting flow generatively as a population of digital twins [43]. What that procedure yields, here and elsewhere, is a mean-field, per-cohort and per-outcome probabilistic digital twin. It is mean-field because no individual record constrains a full trajectory and the flow describes the population average about which individual transitions scatter, per-cohort because the coefficients are learned from a pooled patient population rather than from one subject, per-outcome because each class carries its own equations, and probabilistic because what is integrated forward is the posterior rather than a point estimate. What makes it work is not specific to the aorta but the combination of a sharply timed perturbation (in this case surgery), a readout capturing the response at the relevant scale, and descriptors robust enough that an affine system of ODEs can be learned from a few hundred observations. The binding constraints are whether a macroscopic state exists in which the dynamics are low-order, and whether the observation schedule leaves enough structure between scans for a smoothness assumption to hold. Both are constraints on the observational record rather than on the disease.

While the above constraints inform how a similar digital twin can be built for a different organ once it can be described in terms of reduced-order morphological coordinates, the study framework opens access to many other questions about the aorta itself. Here we restricted the scope to a particular anatomic region (abdominal) and physiological process (post-surgical recovery). The same size–shape metrics are also directly applicable to the *thoracic* aorta where both thoracic aortic aneurysms and dissections can occur; the detailed dynamics of those are different but might be describable in the same dynamical systems language. Dynamical descriptions can be prospectively built for pre-surgical observations of the natural progression of aortic disease. Even prior to the onset of a pathology, aortic size and shape slowly evolve across the human lifetime, dominated by stochasticity rather than deterministic growth [44]. With enough data about those different physiological regimes, digital twins can be extended to a more unified model of health, disease, and recovery.

## Data availability statement

The de-identified extracted morphological features and analysis code supporting this study are available at https://github.com/SurgBioMech/aaa-dynamics. Raw imaging data are not available.

## Ethics approval statement

This retrospective study was approved by the Institutional Review Board of the University of Chicago (IRB IRB20-0653, IRB21-0299) and was conducted in accordance with the Declaration of Helsinki.

## Patient consent statement

A waiver of informed consent was granted by the IRB owing to the retrospective design and the use of de-identified imaging data.

## Permission to reproduce material from other sources

Not applicable. All figures are original to this work.

## Clinical trial registration

Not applicable. This study is not a clinical trial.

## Code and data availability

Code for data processing, model construction, and analysis is available at https://github.com/SurgBioMech/aaa-dynamics, building on the open-source Z–SINDy implementation [45]. The per-patient, per-scan anatomic feature data supporting these findings is available from the same repository.

## Acknowledgments

This work was supported by the National Institutes of Health, USA, NHLBI Grant R01-HL159205 (to L.P.).

## SUPPORTING INFORMATION

### S1 Study cohorts

Table S1 summarizes demographic and clinical metadata for the two centers (Chicago, *n* = 40, and Erasmus, *n* = 60). The centers differ substantially in surveillance cadence: Chicago patients have irregular follow-up over a window extending to eight years, with CT resolutions spanning 0.3–3.0 mm, whereas Erasmus patients contribute exactly two scans (peri-operative and 12-month) at uniform 0.8 mm resolution. These are reported for transparency, and all analyses use the pooled dataset and no covariate adjustment or site stratification was performed.

**Table S1:**
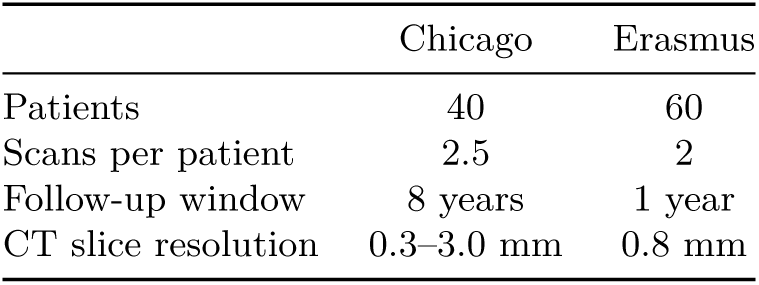
Cohort composition and acquisition characteristics.

### S2 Outlier removal and threshold sensitivity

The derivative-magnitude bounds applied during curation are anisotropic and class-specific rather than a single symmetric window because the two classes carry different prior expectations about which directions of motion are physiological. A regressing sac is, by definition, one whose size is decreasing, so a positive d*A/*d*t* in a regressing patient is far more likely to reflect a segmentation failure than a real event. We accordingly cap regressing transitions at 1.0 yr^−1^ in size and leave them unbounded below so that rapid regression is never filtered out. A stable sac carries no directional expectation, so we apply a symmetric bound of ±3.0 yr^−1^ in both coordinates, i.e., three times the non-pathological normalization scale per year, a rate of change large enough that it describes measurement failure rather than stable morphology. Applied to the whole dataset, 14 of the 120 consecutive real-scan transitions entering this step were flagged and removed, 6 from the Chicago cohort and 8 from Erasmus. Because both endpoints of a flagged transition are removed, a small number of patients drop out of the derivative record entirely at this step, leaving 88 patients with at least one usable derivative.

### S3 Selecting the upsampling step

The interpolated points are not independent observations, and the Z–SINDy posterior covariance **Σ** = *ρ*^2^**C**^−1^ sums over every training row, so its width shrinks as Δ*t* → 0 whether or not the added rows carry new information. A posterior that tightens under interpolation is therefore not by itself evidence of anything, and the choice of Δ*t* has to rest on quantities that are not guaranteed to improve by construction. We refit the model at a sequence of upsampling resolutions from the ground truth CT scans to daily spacing, holding every other choice fixed and using a deliberately non-sparsifying regularization so that all library terms remain active at every resolution. The primary criterion is the stability of the fixed points and the two characteristic timescales of each class (Fig. S1), quantities that must both stop moving with resolution and agree with the steady-state dynamics of just the ground truth, non-upsampled data (“CT”).

Periodic but coarse interpolation is actively harmful, as demonstrated by the results in Fig. S1 together with the coefficient stability and variance explained diagnostics in Figs. S2 and S3. This is the expected failure mode: a grid coarser than the typical inter-scan interval resamples a trajectory at fewer points than it already has, discarding measurements rather than interpolating between them. From Δ*t* ≲ 0.5 yr onward the systems stabilize, with both fixed points and all four timescales varying only mildly from half-year down to daily resolution against the several-fold swings, and one change of sign in the stable shape fixed point, seen at coarser steps. Within that plateau the regressing fixed point sits at (2.00, 1.00) against (1.90, 0.99) from the CT scans, and the faster regressing timescale at 1.3 years against 1.2, so the interpolated models reproduce the reference rather than depart from it. We therefore fixed Δ*t* = 1*/*12 yr for every model reported in this work as it sits well inside the plateau of metric stability.

**Figure S1:**
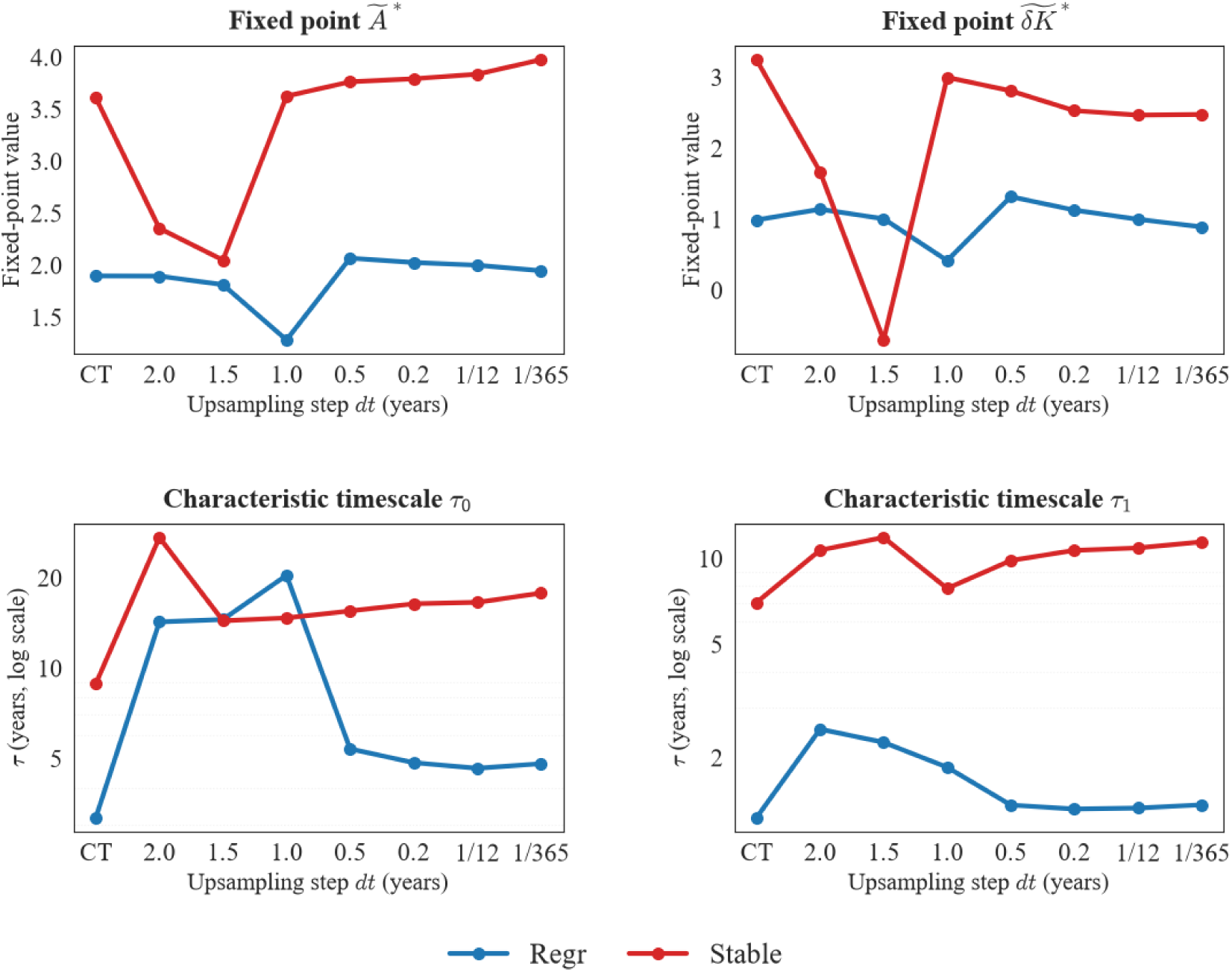
Fixed points and timescales select the interpolation step. Each panel tracks a derived property of the fitted flow against the upsampling step Δ*t*, from just ground truth scans (“CT”, leftmost) to daily spacing. Top row: the two coordinates of the fixed point. Bottom row: the two characteristic timescales *τ* = 1*/*|Re *λ*|, on a log axis. Blue regressing, red stable. Coarse interpolation destabilizes every quantity, most severely between Δ*t* = 2.0 and 1.0 yr. All four quantities plateau below Δ*t* ≈ 0.5 yr and, within that plateau, sit close to the CT values, which are the external reference the interpolated models should reproduce. The chosen upsampling step Δ*t* = 1*/*12 yr lies inside the plateau.

Fig. S2 shows the posterior mean of each of the six coefficients, with error bars giving one posterior standard deviation, for both classes across the resolution sweep. The pattern matches the derived quantities of Fig. S1: the means vary between Δ*t* = 2.0 and 1.0 yr and are stable from 0.5 yr down to daily resolution. The error bars contract monotonically as rows are added, which is expected by construction and is the reason the widths alone cannot select Δ*t*.

Fig. S3 reports variance explained against the CT dataset as a function of Δ*t*. Both classes fall well below the CT baseline at every step of one year or coarser, reaching their minima between Δ*t* = 2.0 and 1.5 yr, and both recover and flatten from 0.5 yr onward. The training-set sizes make the mechanism explicit: resampling onto a two-year grid leaves 24 regressing and 11 stable derivative rows, and a 1.5-year grid leaves 32 and 18, against 46 and 55 from the CT scans themselves. A grid coarser than the typical inter-scan interval is therefore discarding measurements rather than interpolating between them. Within the plateau the regressing model recovers its CT-only value of 0.12, while the stable model settles below it, at 0.06 against 0.10. That the stable class does not fully recover is consistent with its dynamics: with timescales of 11 and 17 years, a stable sac moves very little between consecutive scans, so a larger share of the observed scan-to-scan variation is measurement noise that no smooth flow will explain, and interpolating between two nearly identical states adds rows that are close to uninformative.

**Figure S2:**
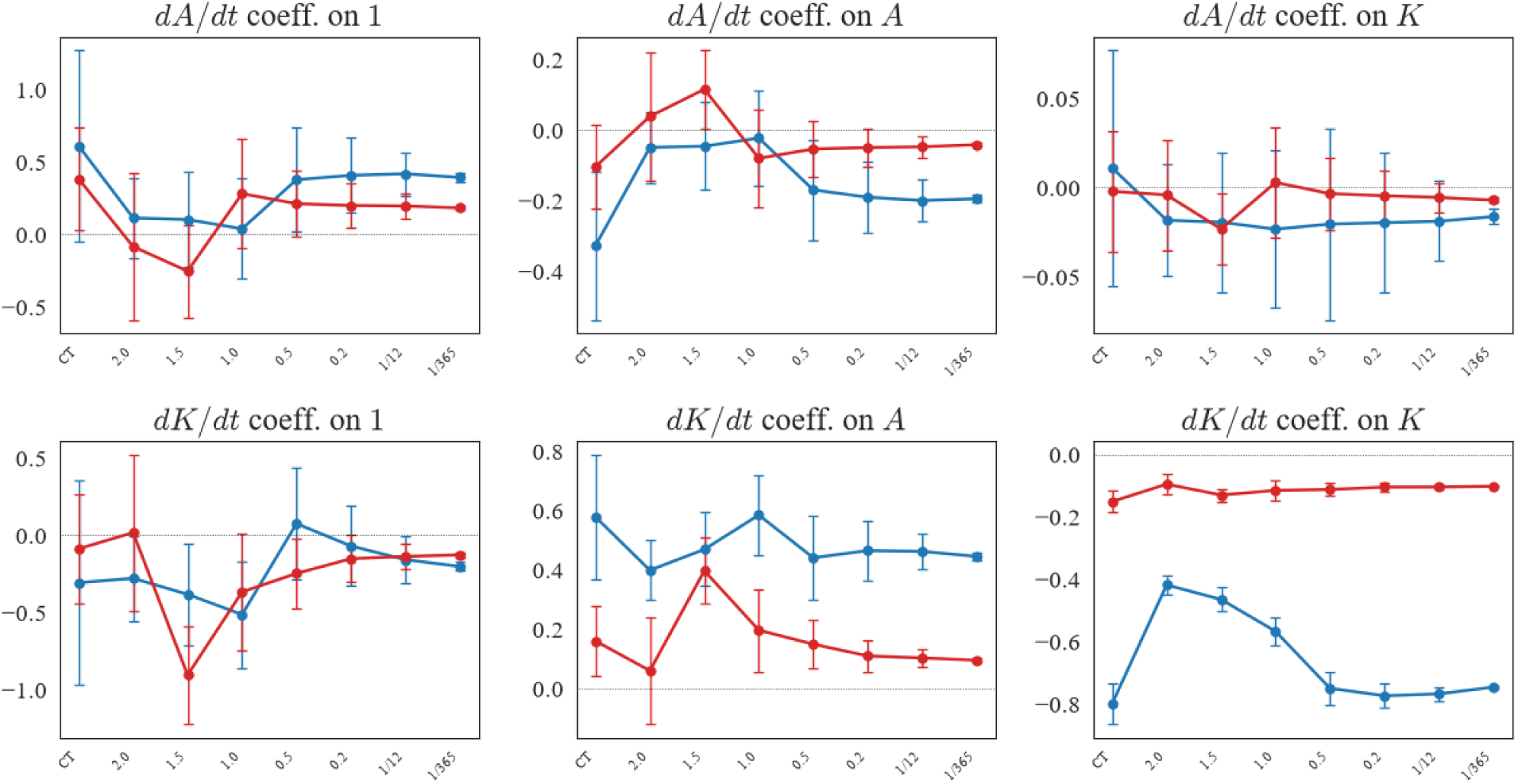
Coefficient stability against upsampling extent. One panel per coefficient of the affine system, with the two rows of the panel grid corresponding to the *Ã* and *δ̃K̃* equations. Markers give the posterior mean and error bars one posterior standard deviation. Blue regressing, red stable. “CT” denotes no interpolation. Means are erratic for coarse steps and flat from Δ*t* ≈ 0.5 yr down to daily resolution; the error bars contract monotonically as interpolated rows are added, which is why posterior width alone cannot be used to select Δ*t*.

### S4 Z–SINDy inference and sparsity

We model **ẋ** (*t*) ≈ **Ξ**^T^**Θ**(**x**(*t*)) with the affine library **Θ**(**x**) = [1*, A, δ̃K̃*]^T^. Coefficients are inferred in a Bayesian framework with a Gaussian likelihood penalizing squared residuals between observed and predicted derivatives,

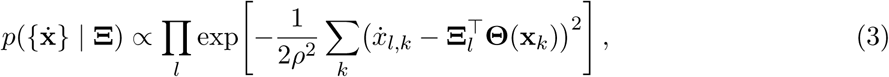

and a sparsity-inducing Bernoulli–Gaussian prior over active-set indicators *γ*, *p*(**Ξ***_l_*) a *_i_*(*δ*(Ξ*_l,i_*) + *w*(Ξ*_l,i_*)*e*^−^*^λn^*), with *λ* controlling parsimony. Conditioning on a fixed active set yields a multivariate Gaussian posterior,

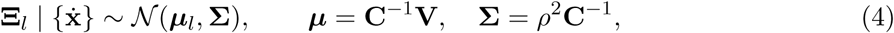

where **C** = ∑*k* **Θ**(**x*k***)**Θ**(**x*k***)^T^ and **V** = ∑*k***Θ**(**x*k***)**ẋ*k*** ^T^. The full posterior is a Gaussian mixture ∑_γ_*p*(**Ξ** | _γ_)Z_γ_, typically dominated by a single *γ*. The resolution parameter is estimated from residual variance, 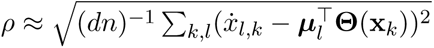.

**Figure S3:**
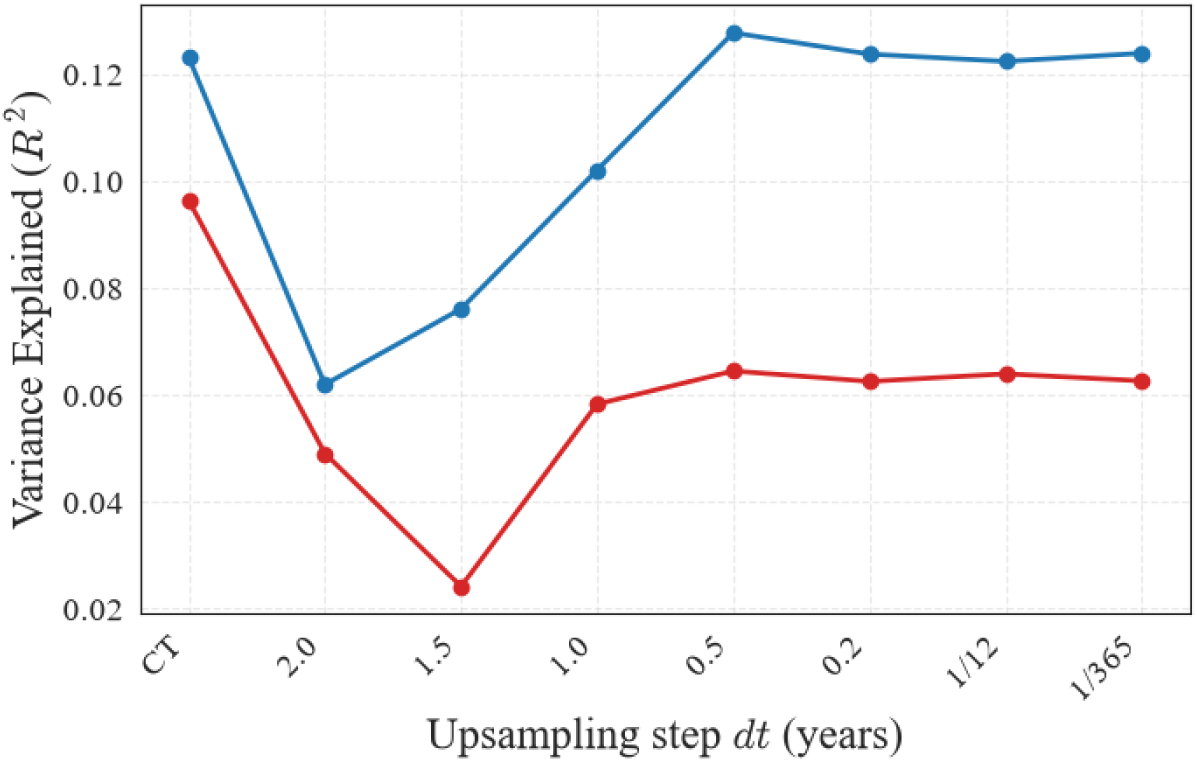
Variance explained against upsampling extent. *R*^2^ scored against the CT scans for each class as a function of Δ*t* (blue regressing, red stable). Coarse interpolation degrades the fit well below the CT baseline, and both curves flatten from Δ*t* ≈ 0.5 yr onward. The regressing model recovers its CT value within the plateau; the stable model settles below it, consistent with a class whose scan-to-scan motion is small relative to measurement noise.

Both **C** and *ρ* sum over the rows of the training set, which is why the interpolation step and the posterior width are coupled: interpolated rows enter the covariance exactly as additional independent observations would, though they are not. Section S3 describes how Δ*t* was chosen so that this coupling does not determine the reported result.

Z–SINDy selects an active set by minimizing a free energy *F* + *λn* in the number of active terms *n*, so negative *λ* rewards additional terms and positive *λ* penalizes them. Sweeping *λ* from −20 to +20 at the canonical upsampling step (Fig. S4), with the number of candidate terms held fixed at three per equation, retains all six coefficients across the entire term-rewarding range and holds in-sample variance explained flat at 0.399 for the regressing class and 0.117 for the stable class. The first terms leave at *λ* = 0, and they are the ones whose posterior means are already near zero, so variance explained falls only marginally as they go, to 0.396 and 0.111 by *λ* = 4, with the stable class losing one further term and settling at 0.099 beyond *λ* = 8. These are in-sample values scored on the upsampled training rows, and are therefore not comparable with the variance explained against the raw CT scans reported in Fig. S3. Sparsification in this problem therefore changes which near-zero coefficients are written as exactly zero and nothing else. We fix *λ* = −5 throughout so that both classes and both preprocessing pipelines are described by the same six-term system and their coefficients can be compared entry by entry.

**Figure S4:**
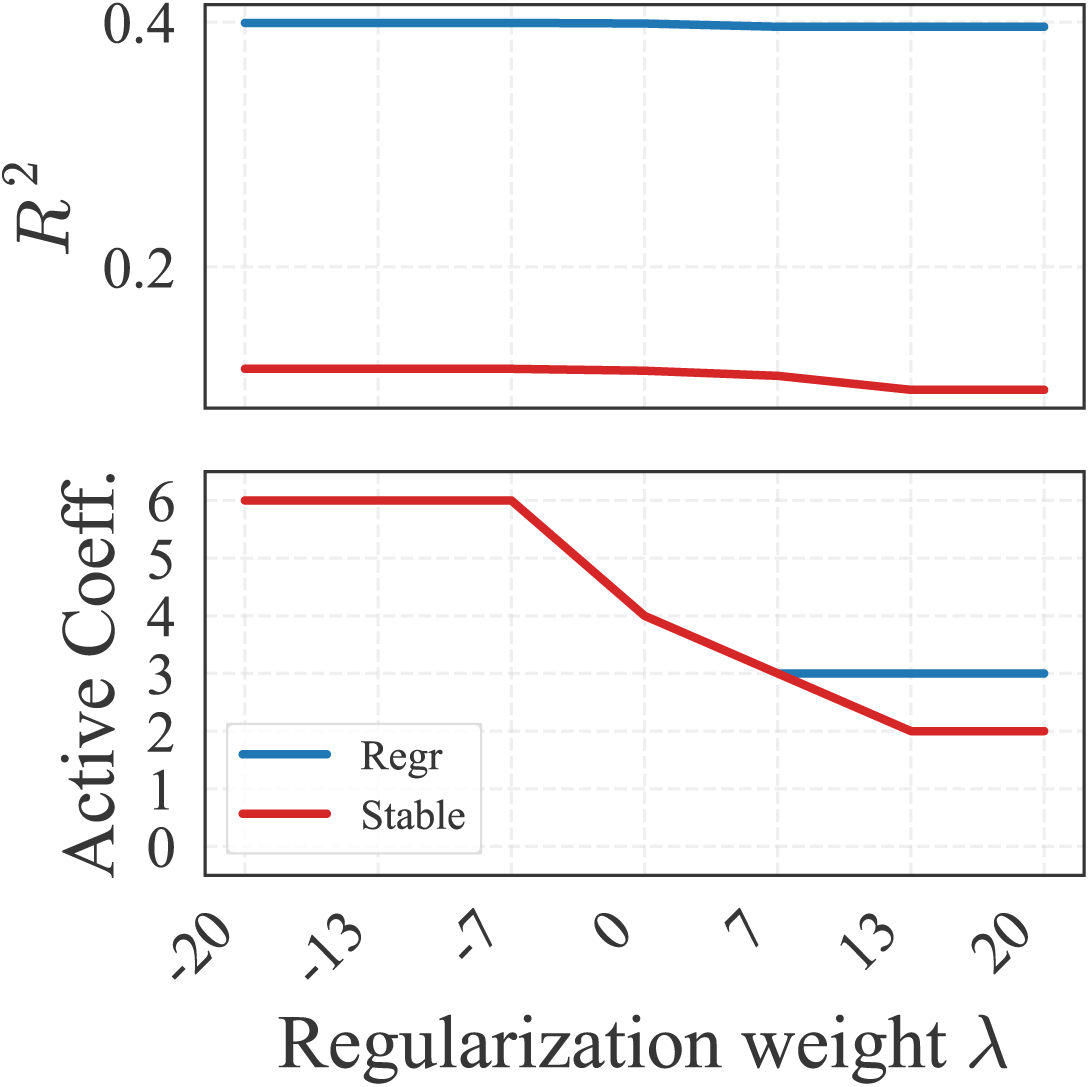
Sparsity sweep at the canonical upsampling step. Top: in-sample variance explained *R*^2^ against the regularization weight *λ* for each class (blue regressing, red stable). Bottom: the number of coefficients the free-energy criterion leaves active in the affine system. Negative *λ* rewards additional terms and positive *λ* penalizes them.

### S5 Bayesian classifiers: likelihoods and normalization

Let **x***_k_* denote the state at *t_k_* and **ẋ** *_k_* its finite-difference derivative. Class-conditional Z–SINDy models are summarized by **Ξ***_c_* and *ρ_c_*.

#### Dynamic classifier

Assuming independent Gaussian errors with variance *ρ*^2^ about the model-predicted derivatives,

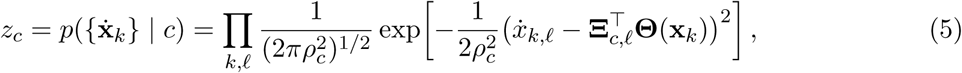

and Bayes’ rule gives *p*(*c* | {**ẋ** *_k_*}) = *z_c_p*(*c*)*/ ∑_c_′ z_c_′ p*(*c*^′^). The model-predicted derivative at *t_k_* is obtained by integrating the class model over [*t_k_* − Δ*t_k_, t_k_*] and differencing, so the comparison is made over the same interval the observed derivative spans.

#### Static classifier

For each class we generate ensembles by sampling initial conditions and coefficients from the posterior and integrating forward, then estimate time-indexed densities *p*(**x** | *c, t*). At time point *t* the classifier scores a patient’s most recent state at or before *t*:

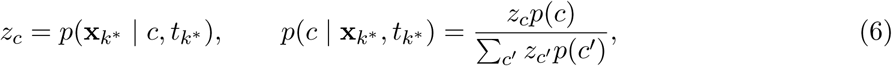

where *k*^∗^ indexes that most recent observation.

#### Evidence windows and eligibility

The two classifiers use deliberately different evidence: the static classifier sees a single most-recent coordinate, the dynamic classifier accumulates every derivative estimate in (0*, t*]. Both share one eligibility rule, in that a patient is classified at *t* only if they have at least one scan at or after *t*, and drops out entirely thereafter. Because the product in Eq. (5) runs over every eligible evidence point, the number of points a patient supplies affects the sharpness of their posterior.

#### Metrics

We report the fraction of eligible patients whose posterior leaves the band [0.1, 0.9] (“classified”) and the accuracy over that subset. Accuracy alone is not interpretable without the classified fraction, since a rule that commits almost never can be perfectly accurate on the few patients it commits to, which is exactly the static classifier’s behavior here.

### S6 Individual-patient classification

The two classifiers were evaluated on the real observational record at 0.5, 1.0, and 1.5 years in two settings. Fig. S5 gives the in-sample evaluation, in which the static densities and the dynamic models are fit once on the full cohort and every patient is scored against them, and Fig. S6 gives the leave-one-patient-out evaluation, in which both are refit for the evaluated patient’s class with that patient wholly excluded, so that every prediction comes from a model that has not seen the individual it is predicting. Training and evaluation both use the upsampled trajectory trained models, so a withheld patient is scored at the same resolution the retained patients were fit at. Numerical values from both appear in Table S2.

The ordering seen on the ensembles (Section 3.5) is reproduced on real patients: the static classifier commits on fewer than one in ten eligible patients at every time point, while the dynamic classifier commits on more than nine in ten. At 0.5, 1.0, and 1.5 years the in-sample static classifier commits on 6.8%, 8.2%, and 6.9% of eligible patients, and is correct on every patient it commits to. The insample dynamic classifier commits on 90.9%, 95.3%, and 96.6% of the same patients, at accuracies of 73.8%, 72.8%, and 53.6%. Leave-one-patient-out validation gives closely similar values, with the static classifier committing on 8.0%, 7.1%, and 6.9% at accuracies of 71.4%, 83.3%, and 100%, and the dynamic classifier on 93.2%, 94.1%, and 93.1% at 72.0%, 72.5%, and 51.9%. That the two settings agree to within a few points indicates the pattern reflects the evidence available to each classifier rather than in-sample optimism.

These two behaviors are the opposite failure modes of the same problem. The static rule almost never commits because the class-conditional state distributions overlap at every early time point, which is the quantitative form of the clinical observation that a single postoperative scan does not indicate what a sac will do. The dynamic rule commits on nearly every patient, which is only possible if the derivative field carries information the state distribution does not. Its accuracy, however, is bounded by how much real derivative evidence each patient supplies. Most contribute a single measured scan pair, so the dynamic posterior is sharp but rests on one chord through state space, and the resulting labels are right about three times in four rather than nine times in ten.

Patient attrition bears on the latest time point. A patient contributes only if their follow-up reached the time point in question, and 88 patients are eligible at 0.5 years and 85 at 1.0 years, but only 29 at 1.5 years, drawn exclusively from the Chicago cohort. The eligible roster is annotated on each row of Figs. S5 and S6, so the three rows are not evaluated on the same population, and the fall in dynamic accuracy at 1.5 years should be read against that change in denominator rather than as a late-time failure of the flow. The scatter panels beside each jitter column show where the classified patients sit in (*Ã, δ̃K̃*). The patients the static rule does commit to are almost all at the extreme upper right of the observed distribution, i.e., the largest and most irregular sacs, which is the region where the two class-conditional densities cease to overlap. This is the direct visual counterpart of the low classified fraction: positional evidence is decisive only for the small minority of patients whose coordinates are atypical.

Read together, these results establish that the learned flows carry class information and generalize to patient trajectories not present in model training, and they mark the ceiling that the observational record imposes: no patient supplies enough measured derivatives for the dynamic rule to be both decisive and highly accurate, and the classifiers cannot be evaluated past eighteen months on more than a third of the cohort. Both limits are properties of the surveillance schedule rather than of the equations, which is what the attrition-aware analysis of Section 3.6 isolates.

**Figure S5:**
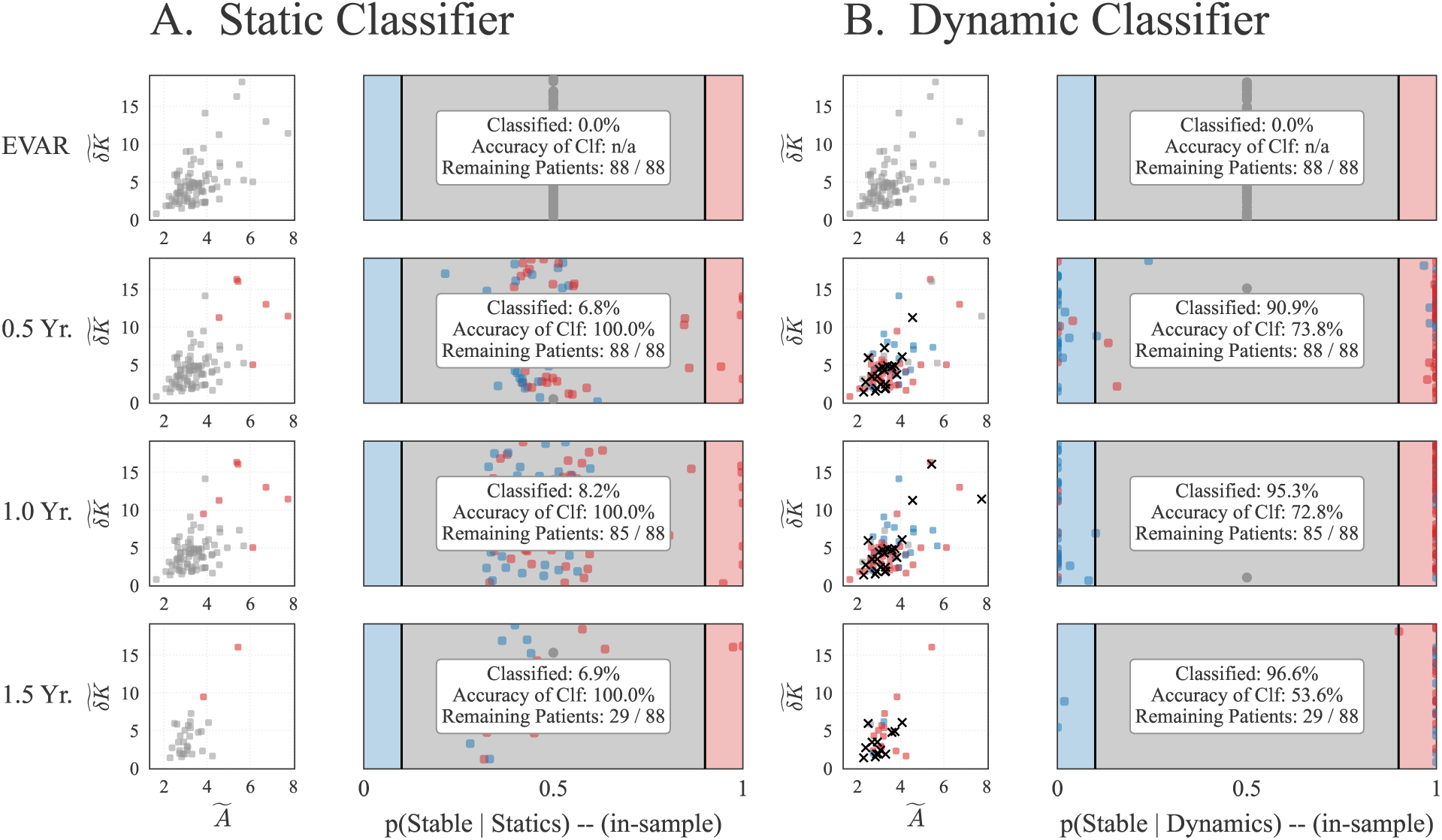
In-sample classification of individual patients. (A) Static classifier. (B) Dynamic classifier. Within each block the left column gives the posterior *p*(stable | ·) for every eligible patient (blue regressing, red stable), with the uncertain band [0.1, 0.9] shaded, and the right column gives the instantaneous (*Ã, δ̃K̃*) scatter of the same patients, with classified patients marked. Rows are index EVAR and 0.5, 1.0, and 1.5 years. The eligible roster is annotated on each row and contracts from 88 of 88 patients to 29 of 88 as follow-up runs out.

**Figure S6:**
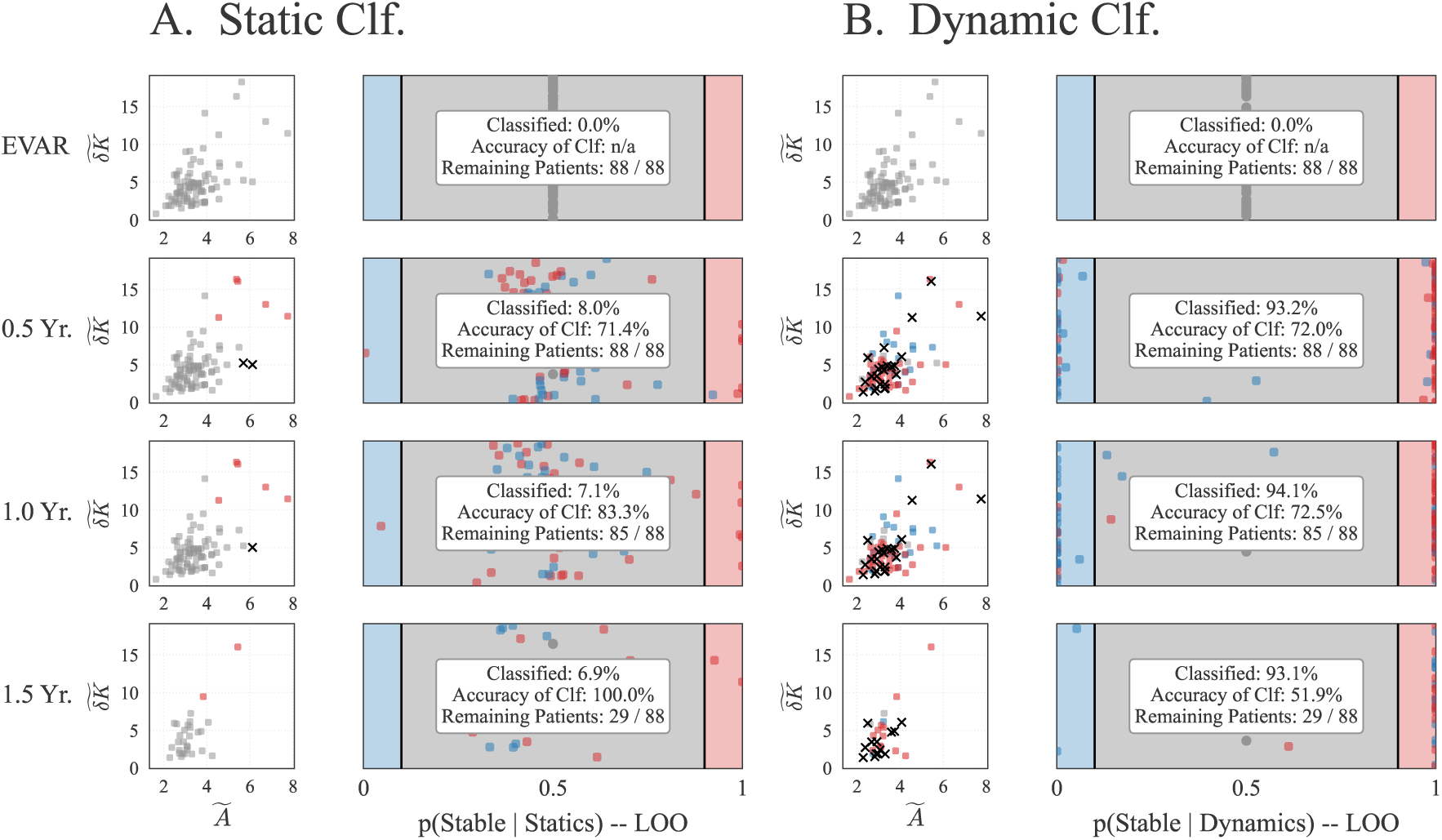
Leave-one-patient-out classification of individual patients. Layout as in Fig. S5, with every patient scored by static densities and a dynamic model refit with that patient wholly excluded. The classified fractions and accuracies agree with the in-sample evaluation to within a few points, indicating that the behavior of each classifier reflects the evidence available to it rather than in-sample optimism.

### S7 Size-only classification and the role of the shape coordinate

Postoperative surveillance is size-based, and the state a clinician acts on is effectively one-dimensional. The normalized fluctuation in integrated Gaussian curvature *δ̃K̃* is what lifts that state to two dimensions here. It is a scale-invariant descriptor by construction [23], i.e., it is not a monotone reparameterization of size, so it carries information about surface irregularity that survives normalization of overall scale [12]. To quantify what the second coordinate buys, we refit the entire pipeline with size alone, using the affine library Θ(*x*) = [1*, A*]^T^ and the scalar equation d*A/*d*t* = *a* + *bA*. Curation, upsampling step, sparsity weight, ensemble construction, and both classifier definitions are held fixed, so the only change is the removal of *δ̃K̃* from the state.

The fitted size-only systems are d*A/*d*t* = 0.446 − 0.229 *A* for regressing sacs and d*A/*d*t* = 0.217 − 0.061 *A* for stable sacs, placing their fixed points at *A*^∗^ = 1.95 and 3.54 with relaxation times of 4.4 and 16.3 years. Both recover the size coordinate of the corresponding two-dimensional fixed point (2.00 and 3.84) and the slower of that model’s two timescales (4.6 and 16.6 years), i.e., the mode that survives the reduction.

Fig. S7 gives the classified fraction for both rules under the size-only model. The dynamic classifier commits on 65.9%, 81.2%, and 82.8% of subjects at 0.5, 1.0, and 1.5 years (58 of 88, 69 of 85, and 24 of 29 eligible patients), against 90.9%, 95.3%, and 96.6% for the two-dimensional model on the same subjects (Table S2), and remains near 80% out to 2.5 years (20 of 25 at two years and 19 of 23 at two and a half). The static classifier commits on 3.4% and 2.4% at 0.5 and 1.0 years, i.e., three patients and then two, and on no subjects at any later time point, against 6.8%, 8.2%, and 6.9% for the two-dimensional model.

Two things follow. First, the ordering is preserved in one dimension, i.e., motion resolves more subjects than position even when the state is size alone, so the trajectory advantage reported in the main text is not an artifact of the extra coordinate. Second, both rules are degraded by the reduction, and the positional rule collapses entirely. The mechanism is visible in the eigenstructure of Table 2. The fast mode of the regressing model, with a relaxation time of 1.3 years, has an eigenvector lying almost along *δ̃K̃*, and a model written in *Ã* alone cannot represent it. What remains is the slower mixed mode at 4.6 years for regressing sacs and 10.9 to 16.6 years for stable sacs, all long relative to the interval over which patients are actually imaged. The size-only static rule fails for the same reason the two-dimensional one does, but more completely, because the *A* marginals are the pair that separate last (Fig. 6, center).

One caveat bounds how much of the drop is informational. The dynamic likelihood of Eq. (5) is a product over coordinates as well as over evidence points, so removing a coordinate halves the number of factors and broadens the posterior mechanically, independent of any information carried by that coordinate. The size-only comparison should therefore be read as the performance of the state a size-based surveillance protocol actually has access to, rather than as an isolated measurement of the information content of *δ̃K̃*.

**Figure S7:**
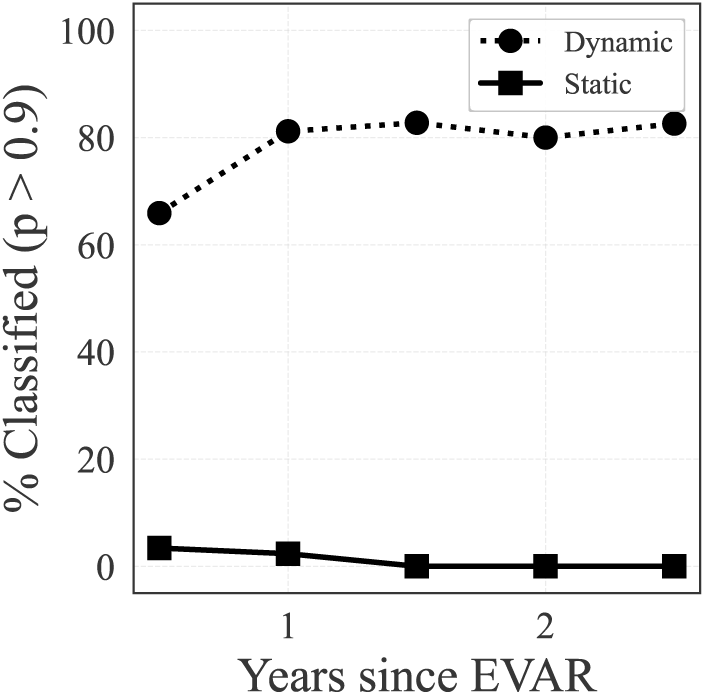
Classification using the size coordinate alone. Percentage of eligible subjects whose posterior leaves the band [0.1, 0.9], as a function of time since repair, for the dynamic (dotted, circles) and static (solid, squares) classifiers built on the one-dimensional interpolated model d*Ã/*d*t* = *a* + *bA*. The dynamic rule retains most of its decisiveness without the shape coordinate, while the static rule commits on three of eighty-eight subjects at half a year and on none beyond 1.5 years. Corresponding two-dimensional values are in Table S2.

### S8 Attrition-aware classification at short horizons

The main-text attrition-aware analysis (Fig. 7) is evaluated at one, two, and five years, which is the horizon over which the class priors move appreciably. Fig. S8 repeats the analysis at 0.5, 1.0, and 1.5 years, matching the time points used for the individual-patient evaluations so that the two can be compared directly. Over this window the prior is nearly static, moving only from *p*(stable) = 0.53 to 0.48, because both classes remain essentially complete through the first year.

The ordering is unchanged and the contrast is sharper at early times. The dynamic classifier commits on 83.2%, 92.3%, and 93.8% of subjects at 0.5, 1.0, and 1.5 years, at accuracies of 99.8%, 99.5%, and 99.4%. The static classifier commits on 3.1%, 41.0%, and 67.5% at accuracies of 87.3%, 91.4%, and 93.3%. At half a year, when the entire enrolled cohort is still under surveillance, the trajectory readout has resolved more than four subjects in five and the positional readout fewer than one in thirty.

**Figure S8:**
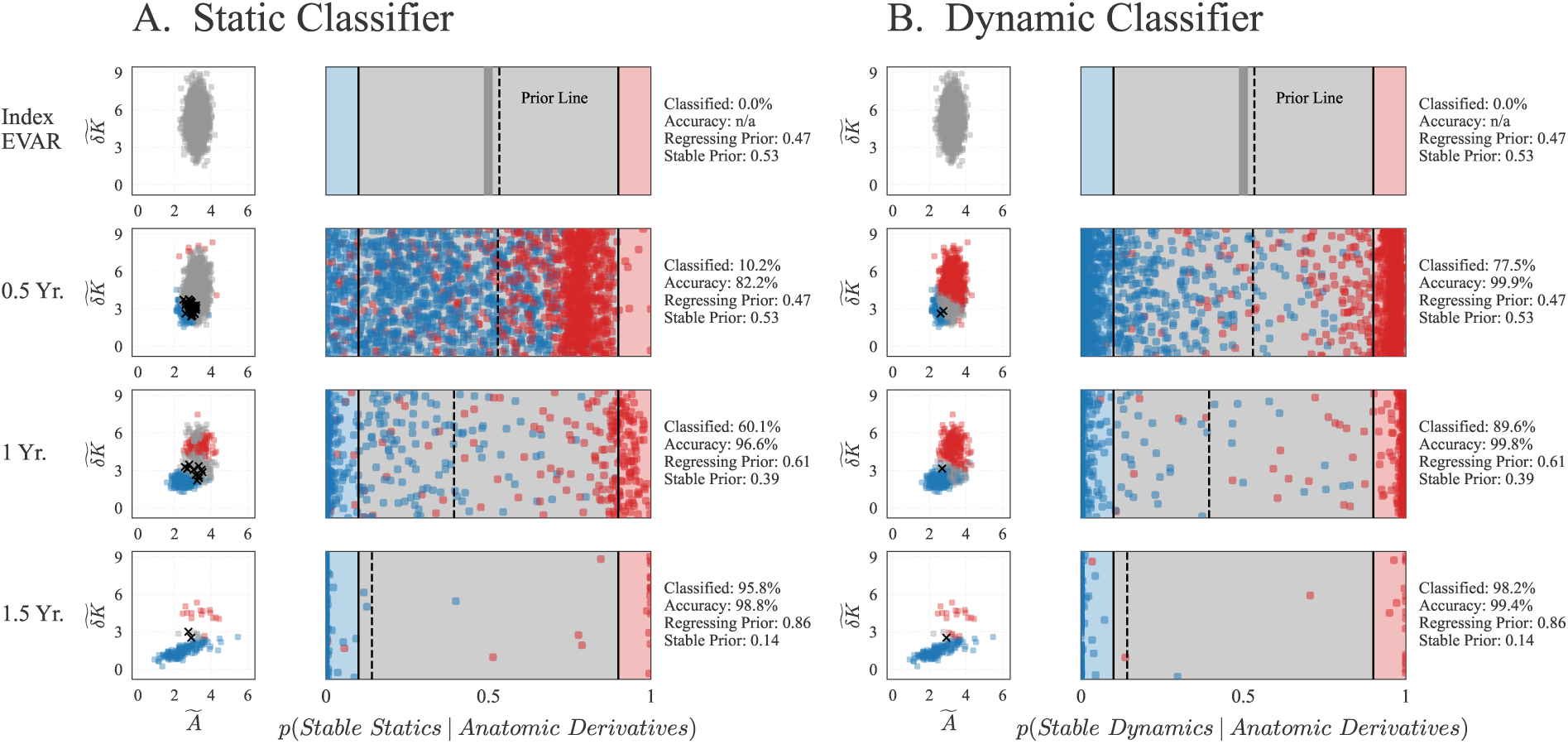
Attrition-aware classification over the first eighteen months. Layout as in Fig. 7, evaluated at index EVAR and 0.5, 1.0, and 1.5 years rather than at one, two, and five. Class priors move only from 0.47*/*0.53 to 0.52*/*0.48 regressing to stable over this window, since both classes remain essentially complete through the first year, so the separation between the two classifiers here reflects the evidence they read rather than the prior they carry.

### S9 Complete classifier results

Table S2 collects every classifier evaluation reported in this work: in-sample, leave-one-patient-out, and size-only on real patients, and on the ensembles under both a fixed prior and attrition-matched dropout with time-dependent priors. Time points differ between the real-patient analyses (0.5, 1.0, and 1.5 years, set by the surveillance record) and the long-horizon ensemble analyses (1, 2, and 5 years, where the horizon is unconstrained), and are labeled accordingly. The attrition-aware analysis is reported at both.

**Table S2:** All Bayesian classifier evaluations. “Classified” is the percentage of eligible subjects whose posterior leaves [0.1, 0.9], and accuracy is computed over that subset. Rows are grouped by the time points at which they are evaluated. The size-only rows use the one-dimensional model of Section S7, and the *p* = 0.5 ensemble rows are the fixed-prior evaluation of Section 3.5. A dash marks an evaluation for which only the classified fraction is reported.

|  |  | Classified (%) |  |  | Accuracy (%) |  |  |
| --- | --- | --- | --- | --- | --- | --- | --- |
| Evaluation | Classifier | $t_1$ | $t_2$ | $t_3$ | $t_1$ | $t_2$ | $t_3$ |
| **Evaluated at 0.5, 1, 1.5 *yr*** |  |  |  |  |  |  |  |
| Real patients, in-sample | Static | 6.8 | 8.2 | 6.9 | 100 | 100 | 100 |
|  | Dynamic | 90.9 | 95.3 | 96.6 | 73.8 | 72.8 | 53.6 |
| Real patients, LOO | Static | 8.0 | 7.1 | 6.9 | 71.4 | 83.3 | 100 |
|  | Dynamic | 93.2 | 94.1 | 93.1 | 72.0 | 72.5 | 51.9 |
| Real patients, size-only | Static | 3.4 | 2.4 | 0.0 | – | – | – |
|  | Dynamic | 65.9 | 81.2 | 82.8 | – | – | – |
| Eligible $n$ (real patients) | | 88 | 85 | 29 | of 88 | | |
| Ensembles, attrition-aware | Static | 3.1 | 41.0 | 67.5 | 87.3 | 91.4 | 93.3 |
|  | Dynamic | 83.2 | 92.3 | 93.8 | 99.8 | 99.5 | 99.4 |
| Contributing $n$ (ensembles, attrition-aware) | | 2000 | 1933 | 689 | of 2000 | | |
| Class prior $p(\text{stable})$, attrition-aware, 0.5/1/1.5 yr | | | | 0.53 / 0.53 / 0.48 | | | |
| **Evaluated at 1, 2, 5 *yr*** |  |  |  |  |  |  |  |
| Ensembles, $p = 0.5$ fixed | Static | 11.9 | 36.9 | 55.7 | – | – | – |
|  | Dynamic | 79.6 | 91.5 | 88.6 | – | – | – |
| Ensembles, attrition-aware | Static | 41.0 | 66.8 | 95.2 | 90.2 | 95.7 | 98.8 |
|  | Dynamic | 92.3 | 91.7 | 96.4 | 99.7 | 99.8 | 98.8 |
| Contributing $n$ (ensembles, attrition-aware) | | 1933 | 587 | 168 | of 2000 | | |
| Class prior $p(\text{stable})$, attrition-aware, 1/2/5 yr | | | | 0.53 / 0.39 / 0.14 | | | |

